# Urban and rural prevalence of tuberculosis in low- and middle-income countries: a systematic review and meta-analysis

**DOI:** 10.1101/2025.09.20.25336166

**Authors:** Seyed Alireza Mortazavi, Nicole A Swartwood, Nanki Singh, Melike Hazal Can, Hening Cui, Do Kyung Ryuk, Katherine C Horton, Nicolas A Menzies, Peter MacPherson

## Abstract

**Background:** Urban and rural settings differ in key determinants of tuberculosis (TB) burden, including transmission dynamics, social and structural determinants, and healthcare access. However, understanding of urban and rural TB burden is limited, hindering implementation of public health interventions to end TB.

**Methods and Findings:** We conducted a systematic review and meta-analysis of urban and rural differences in adult pulmonary TB prevalence in low- and middle-income countries between 1993 and 2024. Bayesian multilevel meta-regression was used to estimate pooled urban-to-rural prevalence ratios (PR) for bacteriologically-confirmed and smear-positive TB overall, and by World Health Organization (WHO) region. We also stratified analysis by survey-level risk of bias and TB screening algorithm, investigated time trends, and evaluated associations with country-level TB incidence and population proportion living in urban areas. To estimate the number of people with prevalent TB in urban and rural areas in study countries, and how these have changed between 2000 and 2023, we fitted a Bayesian multivariate model to WHO incidence and case detection ratio data and combined these estimates with assumptions about the duration of treated and untreated TB and the distribution of urban and rural populations.

We included 46 surveys conducted between 2000 and 2019, encompassing 2,331,775 participants. The pooled urban-to-rural prevalence ratio of bacteriologically-confirmed TB was 1.08 (95% credible interval [CrI]: 0.87-1.33) and was 1.21 (95% CrI: 0.92-1.57) for smear-positive TB. However, there were substantial differences between WHO regions: the African Region had higher urban bacteriologically-confirmed prevalence (PR: 1.29, CrI: 1.01–1.61), while the Western Pacific Region had higher rural prevalence (PR: 0.64, CrI: 0.45–0.89), and burden was broadly similar in the South-East Asia Region (PR: 0.86, 95% CrI: 0.64-1.10). Time trends indicated a small increase in the overall bacteriologically-confirmed urban-to-rural prevalence ratio between 2000 and 2019, with a mean PR increase of 2.2% (95% CrI: -2.5 to 8.0%) per year.

We estimated that, for 2023 in the 26 represented study countries (combined population: 2.2 billion urban, 2.4 billion rural), 47% (95% CrI: 34-62%; 6.5 million, 95% CrI: 3.6-11.8 million) of prevalent TB was in urban areas, and 53% (95% CrI: 38-66%; 7.3 million, 95% CrI: 4.2-12.8 million) in rural areas. Within countries, there were striking changes in the urban and rural distribution between 2000 and 2023, with the share of urban cases increasing in nearly all countries.

**Conclusion:** Between 2000 and 2023, TB epidemics have become increasingly urbanised, both in proportional and absolute terms, although with considerable variation across countries and regions. Public health approaches tailored to urban and rural TB epidemiology and demography will be required to end TB.

## Background

Tuberculosis (TB) remains a leading infectious cause of death worldwide, with incidence and mortality concentrated in low- and middle-income countries (LMICs) [1,2]. In 2014, the World Health Organization (WHO) set an ambitious goal to end TB by 2035, yet substantial progress is needed to meet this goal [1].

Understanding variation in TB burden both between and within countries and regions is essential for developing effective public health strategies to address TB determinants and accelerate TB care and prevention [3]. Urban and rural environments differ substantially in the factors that shape TB transmission, healthcare access, and health-seeking behaviours [4]. Urban areas may experience higher TB prevalence due to greater population density, higher rates of HIV infection, and increased exposure to crowded congregate settings and poor outdoor air quality [5]. In contrast, people living in rural areas can face barriers such as limited healthcare access, delayed diagnosis, treatment challenges as well as higher rates of indoor air pollution [6,7]. For instance, in India, whilst 72% of the population lives in rural areas, only 40% of healthcare workers are stationed there [8]. TB epidemiology may also be affected by high rates of recent rural-to-urban migration [9], with rural populations experiencing more rapid aging [10].

Although some national TB prevalence surveys have found differences in TB prevalence between urban and rural areas (e.g. [11]), the magnitude of this difference has not been quantified, and there is little understanding of country and regional patterns and determinants.

Therefore, we conducted a systematic review and meta-analysis using data from national and sub-national TB prevalence surveys conducted in LMICs. We compared urban and rural TB prevalence and estimated burden and secular trends of urban and rural TB within LMICs and WHO regions. We also investigated key determinants to inform public health approaches to TB elimination.

## Methods

We conducted a systematic review and meta-analysis of differences in TB prevalence between urban and rural areas, reporting methods and results in accordance with PRISMA guidelines [12]. Our protocol was prospectively registered (PROSPERO number: CRD42024503853) [13].

### Inclusion and exclusion criteria

We included studies published in English between 1st January 1993 and 1st January 2024 that reported the prevalence of pulmonary TB in both rural and urban adult populations (15 years or older) in LMICs (as defined by World Bank country groupings [14]). We included pre-intervention surveys conducted as part of community cluster randomised trials but excluded post-intervention surveys. Eligible studies reported prevalence estimates—defined as numerator and denominator counts, or unadjusted or adjusted prevalence rates—stratified by urban and rural populations, or provided sufficient data for their calculation. We excluded studies that reported only extra-pulmonary TB or TB prevalence exclusively among children under the age of 15 years. Studies that included data only for symptomatic or healthcare-seeking individuals, such as studies of case notifications, were excluded. We also excluded studies conducted in congregate settings, including prisons, universities, and health facilities. Studies published in languages other than English were excluded due to limited translation resources. Full inclusion and exclusion eligibility criteria are provided in S1 Table.

### Search Strategy

Studies were identified by searching PubMed, Embase, Global Health, and the Cochrane Library. Search terms were defined to identify all relevant studies by utilizing a combination of MeSH terms and keywords related to 1) tuberculosis, 2) prevalence and 3) low- and middle-income countries (LMICs) (S2 Table). We additionally considered all studies from a previous systematic review of sex differences in TB prevalence [15], WHO reports of prevalence surveys sourced from the Global TB Report, as well as the abstracts of the 2023 Union World Conference on Lung Health.

This systematic review was managed using Covidence [16]. After removal of duplicates, titles and abstracts were independently reviewed for relevance by pairs of reviewers, allocated randomly from a pool of eight team members with resolution of discrepancies by a third reviewer or team discussion. Studies included for full-text review were independently assessed against inclusion and exclusion criteria by two reviewers, randomly allocated in pairs from a team of eight, with recording of reasons for exclusion.

Extraction was done in duplicate by pairs of reviewers using a pre-tested form. Data extracted included: title and identification information; survey methodology; and screening methods; case definitions; and prevalence survey results. A consensus reviewer, allocated at random, resolved discrepancies in data extraction, with further group discussion, if required.

### Definitions

We defined bacteriologically-confirmed TB in a study participant as at least one positive sputum result for *Mycobacterium tuberculosis* on testing by smear microscopy, culture, or molecular testing (e.g., Xpert MTB/RIF). Smear-positive TB was defined by a positive smear microscopy result, regardless of other results. Where study definitions varied (for example, requiring more than one positive test to classify a case as bacteriologically confirmed TB), we adhered to the definitions used in each study. We used study definitions of “urban” and “rural”. If a study reported more than two urbanicity categories, but these categories could be grouped into urban and rural, they were aggregated accordingly (e.g., “state-urban” and “regional-urban” were grouped as urban). If a study reported an urbanicity category that could not be clearly classified as urban or rural, data for this category were excluded.

### Outcomes

The primary outcome was the urban-to-rural TB prevalence ratio, defined as the ratio of bacteriologically-confirmed TB prevalence in urban versus rural areas. In secondary analysis we estimated variation in ratios stratified by WHO global region and survey country, and the urban-to-rural prevalence ratio of smear-positive TB. We also conducted meta-regression to investigate predictors urban-to-rural prevalence ratio (overall, and by WHO region). Potential predictors included the proportion of the study country population who resided in urban areas [17] and country-level TB incidence in the prevalence survey year [1]. Finally, we constructed epidemiological models to estimate the numbers of people with prevalent TB in urban and rural areas, and how these have changed over time.

### Risk of bias assessment

Risk of bias was assessed using a tool developed by Hoy *et al*. for prevalence surveys [18]. Two reviewers each independently classified studies as low, medium, or high risk of bias across domain questions and made an overall risk of bias assessment (high, moderate, low, unknown), with discrepancies resolved by a third consensus reviewer.

### Statistical analysis

We summarised study characteristics, including setting, survey methodologies, TB screening and diagnosis methods, and case definitions. In initial descriptive analysis, we extracted or calculated (where not reported) the crude urban and rural prevalence for each study, based on the reported number of prevalent TB cases and population denominators, or rates, and calculated the crude urban-to-rural prevalence ratio. Because prevalence surveys are usually conducted within clusters, the reported 95% confidence intervals are often adjusted for clustering, typically using robust standard errors or regression models [19]. Where available, we extracted these cluster-adjusted confidence intervals. Moreover, typically in prevalence surveys certain population groups, usually men and younger people, are under-represented and surveys often undertake weighted analysis to adjust prevalence estimates for this under-representation [19]. Where reported, we additionally extracted adjusted urban and rural TB prevalence estimates from a WHO-recommended regression approach, which uses multiple imputation and inverse probability weighting to address sampling bias [19]. To pool estimates of the urban-to-rural prevalence ratios, we preferentially used survey point estimates and 95% confidence intervals based on adjusted/imputed prevalence rates, followed by crude prevalence rates, and using counts of numerators and denominators where neither prevalence rate was available (data hierarchy shown in S3 Fig.).

We constructed hierarchical Bayesian meta-regression models [20] to estimate the pooled log-odds of the urban-to-rural prevalence ratio for bacteriologically-confirmed and smear-positive pulmonary TB, weighted for precision by the log of the study-specific prevalence ratio standard error, and with a random-effects grouping term for each prevalence survey and country (S4 Text). Subsequent meta-regression models included terms for WHO region, estimated country TB incidence in the prevalence survey year [1], survey end year, the percentage of country population estimated to be living in urban areas [17], whether symptom screening was used to identify participants for sputum testing, and risk of bias. Priors were weakly informative. We retained 4000 posterior draws for each parameter from the posterior and exponentiated and summarised these results to obtain mean posterior prevalence ratios and 95% credible intervals (CrI).

To estimate the number and percentage of people with prevalent TB in urban and rural areas in study countries, and how these have changed between 2000 and 2023, we fitted a Bayesian multivariate model to WHO incidence and case detection ratio data, and combined these estimates with assumptions about the duration of treated and untreated TB (stratified by smear status) [21] and the distribution of urban and rural populations (S4 Text) [17].

Models were fit using the brms interface (version 2.22.0) [22] to Stan (version 2.37.0) [23]. We assessed model convergence using 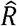 statistics, effective sample size measures, trace plots of chains, and posterior predictive plots. Model predictions and summaries used the tidyverse [24] and tidybayes [25]. All analyses were conducted using R version 4.5.0 [26]. The extracted datasets, R codes and additional material are publicly accessible at https://github.com/nswartwo/tb-lmic-prevalence-review/.

## Results

Our search identified 7,716 records, of which 7,077 were screened after duplicate removal. Following abstract screening, 212 studies underwent full-text review, and 45 studies reported TB prevalence disaggregated by urban and rural setting and were included in this analysis [11,27–70]. One study in China reported two separate prevalence surveys [68], giving a total of 46 prevalence surveys that contributed to quantitative analysis (Figure 1).

**Figure 1:**
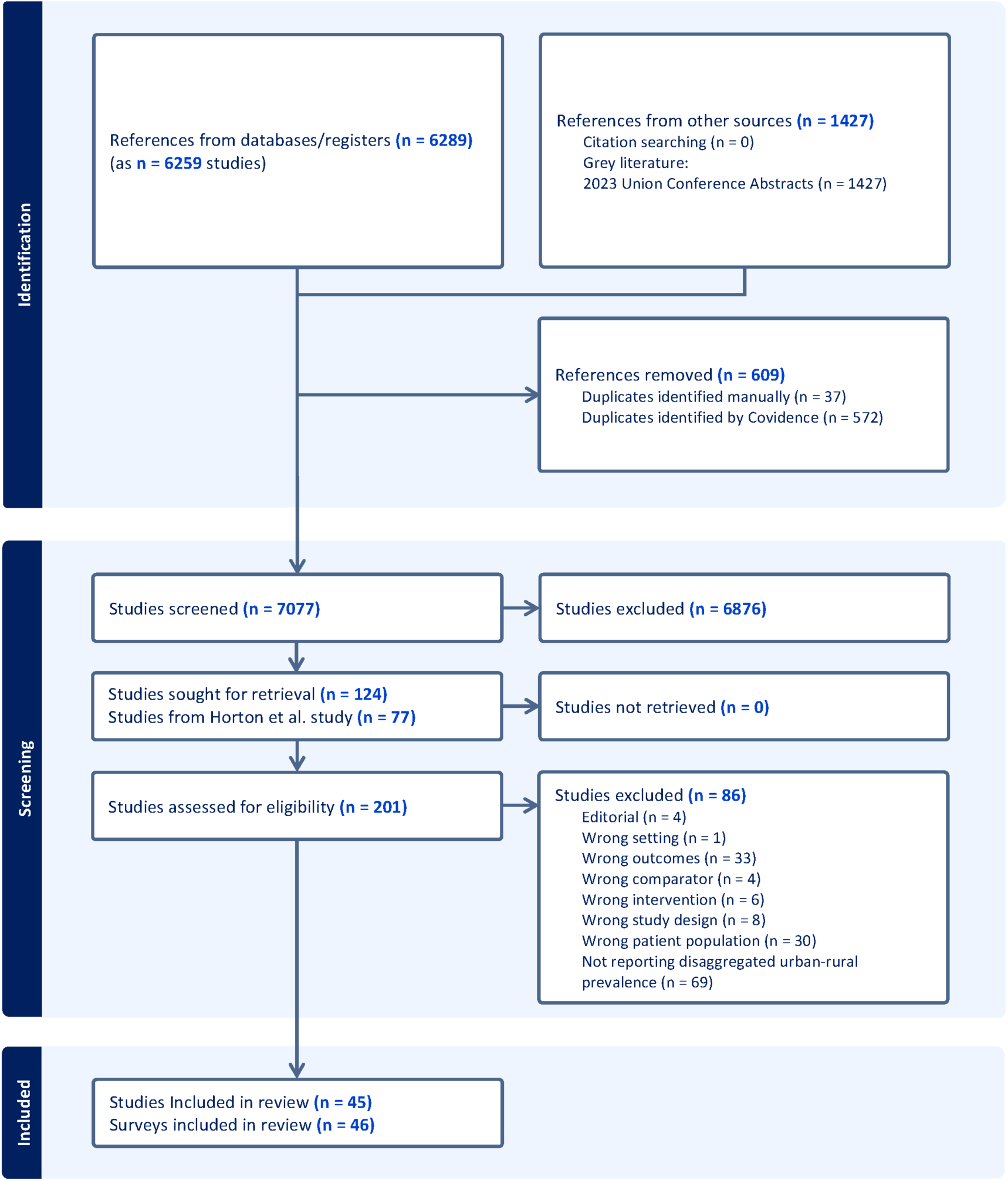
PRISMA Flow Chart. Some studies included results from more than one prevalence survey and other surveys were described in multiple studies

### Study characteristics

The 46 included prevalence surveys were conducted between 2000 and 2019 across 26 countries from four WHO regions. The African (n=19) [27,29–31,34,41,43–50,52,53,57,63,66] and South-East Asia (n=14) [28,35–37,42,51,55,56,59,61,62,64,65,69] Regions contributed the largest number of surveys, followed by the Western Pacific Region (n=11) [11,32,38–40,54,58,67,68,70] and the Eastern Mediterranean Region (n=2) [33,60]. India (n=6) [28,37,59,61,62,64], China (n=5) [11,39,68,70], and Ethiopia (n=4) [30,34,41,66] had the greatest number of surveys among individual countries. Overall, 32/46 (69.6%) of surveys were nationally representative surveys, most of which used multistage cluster sampling. Details of included studies are provided in S5 Table.

In 36/46 (78.3%) studies, participants were eligible for sputum testing if they reported TB symptoms or had abnormalities on chest radiography, eight (17.4%) screened with symptom screening alone, and the remaining two (4.3%) collected sputum samples from all participants without a screening stage.

Across the 46 studies, 2,518,863 individuals were eligible for screening (S6 Table). Screening eligibility was not always reported stratified by urban or rural status, but where reported, 39.9% of screening-eligible participants were from urban settings, and 60.1% were from rural settings. Overall, 92.6% of eligible individuals participated in screening, with 38.5% from urban settings and 61.5% from rural settings, where reported. Bacteriologically-confirmed TB was diagnosed in 7,141 participants (where reported: urban: 39.9%; rural: 60.1%). Among these, 3,213 had smear-positive TB (where reported: urban: 33.2%; rural: 66.8%) and 3,765 had culture-positive TB (where reported: urban: 36.3%; rural: 63.7%).

### Urban-to-rural prevalence of bacteriologically-confirmed TB

Across all 46 surveys, reported crude urban-to-rural prevalence ratios ranged from 0.20 (China, Shandong Province, 2010) to 2.71 (Malawi National TB Prevalence Survey, 2013). The lowest crude ratios—below 0.5—were reported in eight surveys, all conducted in China and India. In contrast, all surveys reporting urban-to-rural prevalence ratios greater than two were from the WHO African Region. Overall, 24/46 (52.2%) of surveys reported a crude urban-to-rural ratio greater than one.

In our main model, which excluded two studies from the Eastern Mediterranean Region with disparate crude estimates, the pooled urban-to-rural prevalence ratio for bacteriologically-confirmed TB was 1.08, (95% CrI: 0.87–1.33). The urban-to-rural prevalence ratio differed substantially between WHO regions (Figure 2). The African Region had higher prevalence in urban areas (PR: 1.29, 95% CrI: 1.01–1.61), whereas the Western Pacific Region had higher prevalence in rural areas (PR: 0.64, 95% CrI: 0.45–0.89) and was broadly similar in the South-East Asia Region (PR: 0.86, 95% CrI: 0.64–1.10).

**Figure 2:**
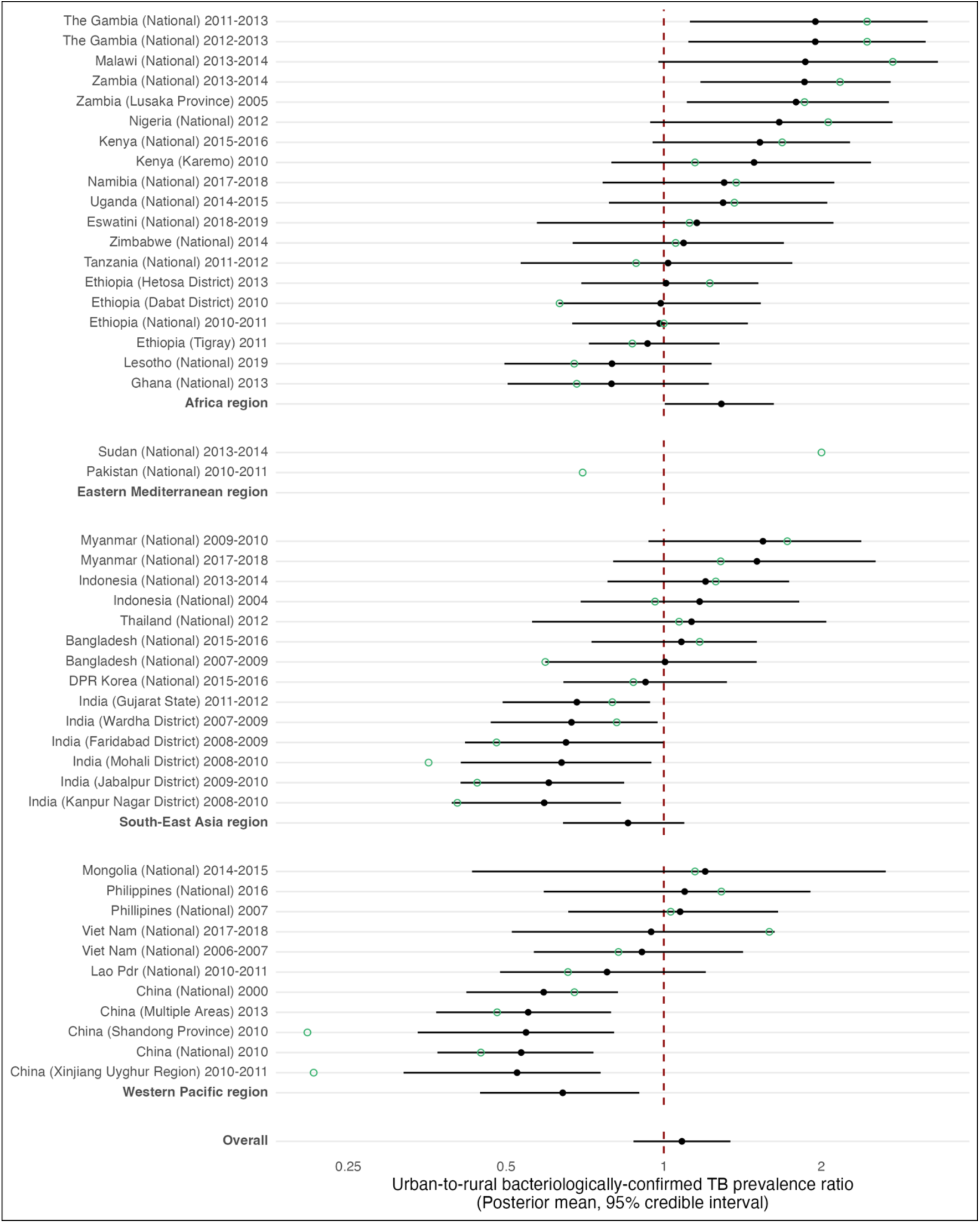
Forest plot of urban-to-rural prevalence ratios of bacteriologically-confirmed tuberculosis. Green points are crude survey urban-to-rural prevalence ratios, and black points and error bars posterior means and 95% credible intervals from multilevel meta-analysis models. Only empirical estimates are shown for Eastern Mediterranean Region as these were not included in multilevel meta-analysis model. Overall pooled estimate includes data from African Region, South-East Asia Region, and Western Pacific Region, but not Eastern Mediterranean Region.

### Change in the urban-to-rural prevalence ratio over time

Time trend analyses were consistent with a small increase in the overall urban-to-rural prevalence ratio of bacteriologically-confirmed TB between 2000 and 2019, with a mean increase of 2.2% (95% CrI: -2.5 - 8.0%) per year (Figure 3). By 2019, there was an 86.4% probability that the urban-to-rural prevalence ratio exceeded one. Estimates of the average annual change in the PR within individual WHO regions were imprecise: African Region (-1.32, -9.0 to 6.6%); South-East Asia Region (2.9%, -4.8 - 12.6%); Western Pacific (-1.8%, -11.9 - 7.4%). By 2019, the posterior probability that the urban-to-rural prevalence ratio exceeded one was 74.6% in the African Region, 56.0% in the South-East Asia Region, and 9.4% in the Western Pacific Region.

**Figure 3:**
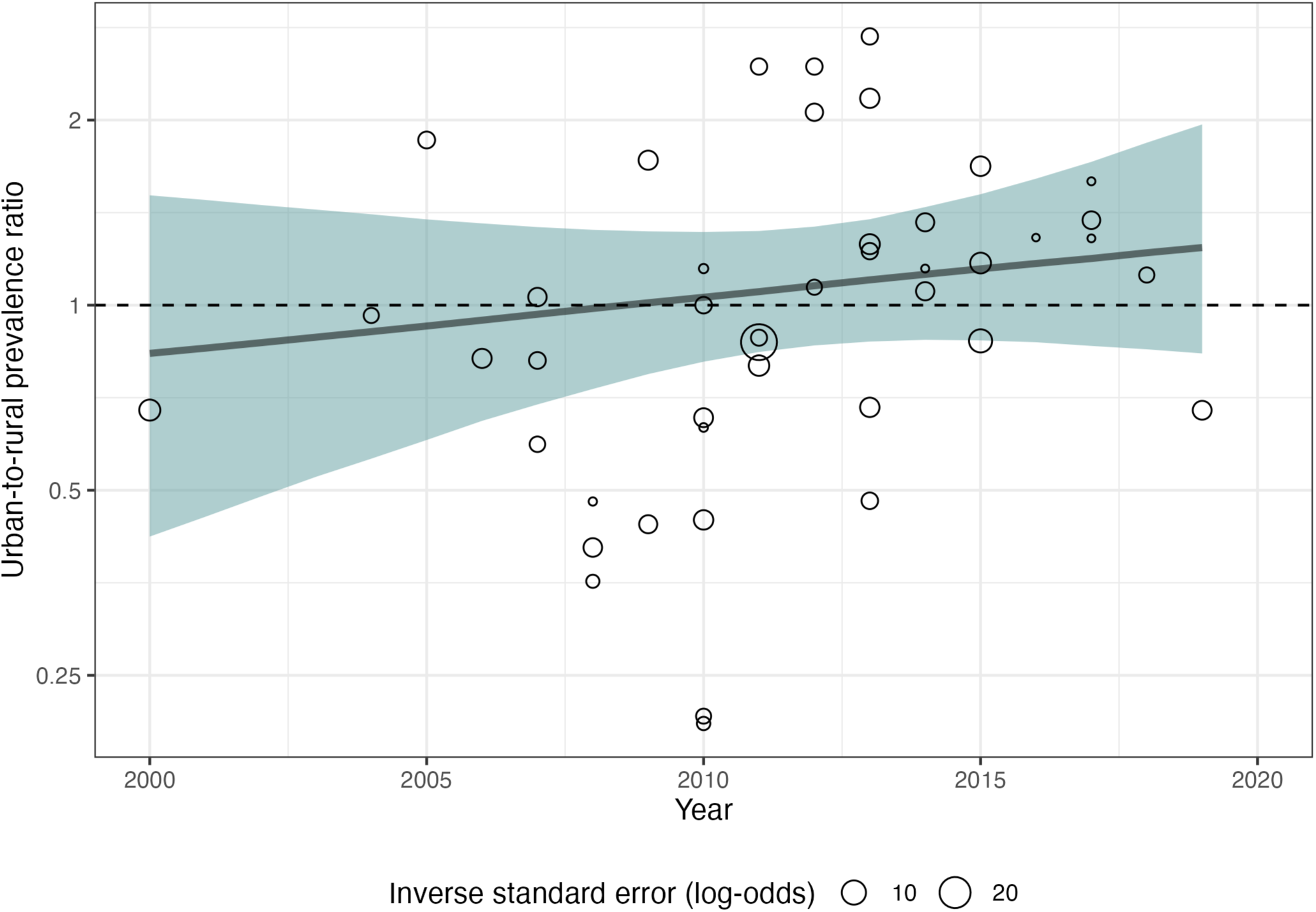
Time trend in the urban-to-rural prevalence ratio for bacteriologically-confirmed pulmonary tuberculosis: 2000-2019. In univariate analysis, among the 44 surveys in 24 LMICs in the African, South-East Asia, and Western Pacific Regions, there was evidence that nationally-representative prevalence surveys had higher urban-to-rural prevalence ratios than sub-national surveys (multiplicative effect: 1.70, 95% CrI: 1.00-2.76) (Table 2). Although this association was somewhat attenuated in multivariable analysis, the magnitude and directionality of associations were consistent across all three global regions. Associations estimated for other predictors were consistently weak.

**Table 1.** Multiplicative effect of covariates on urban-to-rural prevalence ratio of bacteriologically-confirmed TB prevalence.

|  | Global (24 LMICs) |  | African Region |  | South-East Asia Region |  | Western Pacific Region |  |
| --- | --- | --- | --- | --- | --- | --- | --- | --- |
|  | Univariate | Multivariable | Univariate | Multivariable | Univariate | Multivariable | Univariate | Multivariable |
| Risk of bias (not low vs low) | 1.09<br>(0.61-1.85) | 1.12<br>(0.57-2.30) | 1.04<br>(0.64-1.61) | 1.07<br>(0.62-1.77) | 1.07<br>(0.62-1.73) | 1.03<br>(0.48-1.90) | 1.14<br>(0.64-2.00) | 1.25<br>(0.69-2.95) |
| National TB incidence<sup>†</sup> (per 100/100,000 increase) | 1.10<br>(0.88-1.38) | 1.05<br>(0.84-1.33) | 1.03<br>(0.88-1.20) | 1.03<br>(0.89-1.19) | 1.11<br>(0.93-1.34) | 1.05<br>(0.87-1.29) | 1.15<br>(0.95-1.40) | 1.06<br>(0.88-1.30) |
| Nationally representative prevalence survey (Yes vs No) | 1.70<br>(1.00-2.76) | 1.69<br>(0.92-3.09) | 1.57<br>(0.96-2.38) | 1.64<br>(0.90-2.71) | 1.75<br>(1.14-2.62) | 1.80<br>(1.10-3.14) | 1.73<br>(1.00-2.94) | 1.67<br>(0.89-2.97) |
| Percent of population in urban areas<sup>†</sup> (per % point increase) | 1.00<br>(0.97-1.04) | 0.99<br>(0.95-1.03) | 1.01<br>(0.99-1.02) | 1.00<br>(0.99-1.02) | 1.00<br>(0.99-1.02) | 0.99<br>(0.97-1.01) | 0.99<br>(0.97-1.01) | 0.99<br>(0.96-1.01) |
| Symptom screening used for sputum eligibility (Yes vs No) | 0.99<br>(0.34-2.24) | 1.10<br>(0.34-3.29) | 1.06<br>(0.42-2.23) | 1.08<br>(0.38-3.10) | 1.00<br>(0.39-2.19) | 1.18<br>(0.41-3.36) | 0.89<br>(0.27-2.22) | 1.03<br>(0.26-3.50) |

Eastern Mediterranean Region countries not included. Multivariable models adjusted for all covariates. LMICs: low- and middle-income countries.
<sup>†</sup>In the year in which the prevalence survey was conducted.

### Urban-to-rural prevalence of smear-positive TB

Thirty-one surveys with 1,433,666 total participants (36.7% urban) reported urban-rural smear-positive TB prevalence. The pooled urban-to-rural smear-positive TB prevalence was 1.21 (95% CrI: 0.92–1.57). The estimated smear-positive prevalence ratio was 1.43 (95% CrI: 1.01–1.96) in the WHO African Region, 1.02 (95% CrI: 0.67–1.46) in South-East Asia Region, and 0.93 (95% CrI: 0.51–1.41) in the Western Pacific Region. Overall, time trends were more pronounced than for bacteriologically-confirmed TB; there was an estimated 12.0% (95% CrI: 1.0-24.1%) annual increase in the urban-to-rural prevalence ratio of smear-positive TB between 2000 and 2019, with a 99.5% posterior probability that the prevalence ratio was greater than one in 2019.

### Burden of bacteriologically-confirmed TB in urban and rural populations

Between 2000 and 2023, urban and rural populations increased substantially in the 26 countries represented in the review (S7 Fig.). In some countries, especially China, Indonesia, and Thailand, there was rapid urbanisation (S8 Fig.). By 2023, we estimated that there were 4.60 billion people in these 26 study countries, with 2.20 billion (47.9%) in urban areas, and 2.40 billion (52.1%) in rural areas.

Multivariate modelled estimates of WHO TB incidence and case detection ratio data between 2000 and 2023 are shown in S9 Fig. and S10 Fig., and country-specific estimates of the urban-to-rural prevalence ratio of bacteriologically-confirmed TB in S11 Fig. All countries had declining TB incidence, increasing case detection (excluding in a small number of countries during the COVID-19 pandemic in 2020-2021), and an increasing urban-to-rural prevalence ratio over time.

By modelling changes in TB incidence and case detection data, and incorporating country-specific population structure and urban-to-rural prevalence ratio estimates, we estimated that in these 26 countries in 2023 there were 13.8 million (95% CrI: 8.7-23,2 million) people with prevalent TB. Of these, 47% (6.5 million, 95% CrI: 3.6-11.8 million) were in urban areas, and 53% (7.2 million, 95% CrI: 4.2-12.8 million) were in rural areas. In absolute terms, in 2023, India, Pakistan and Indonesia had the greatest numbers of both urban and rural people with prevalent TB in 2023 (S12 Table). Within the WHO African Region the greatest number of people with prevalent TB in rural areas were in Nigeria, Ethiopia, and Tanzania, whereas the greatest number in urban areas were in Nigeria, Tanzania, and Kenya.

Between 2000 and 2023, there were striking changes in the distribution of TB between urban and rural populations within countries, both in percentage terms (Figure 5), and by absolute numbers (S13 Fig). In all countries, the percentage of people with prevalent TB in rural areas declined, and in urban areas increased. By 2023, the median percentage of prevalent cases was higher in urban than in rural areas in 12 of the 26 countries (Figure 4).

**Figure 4:**
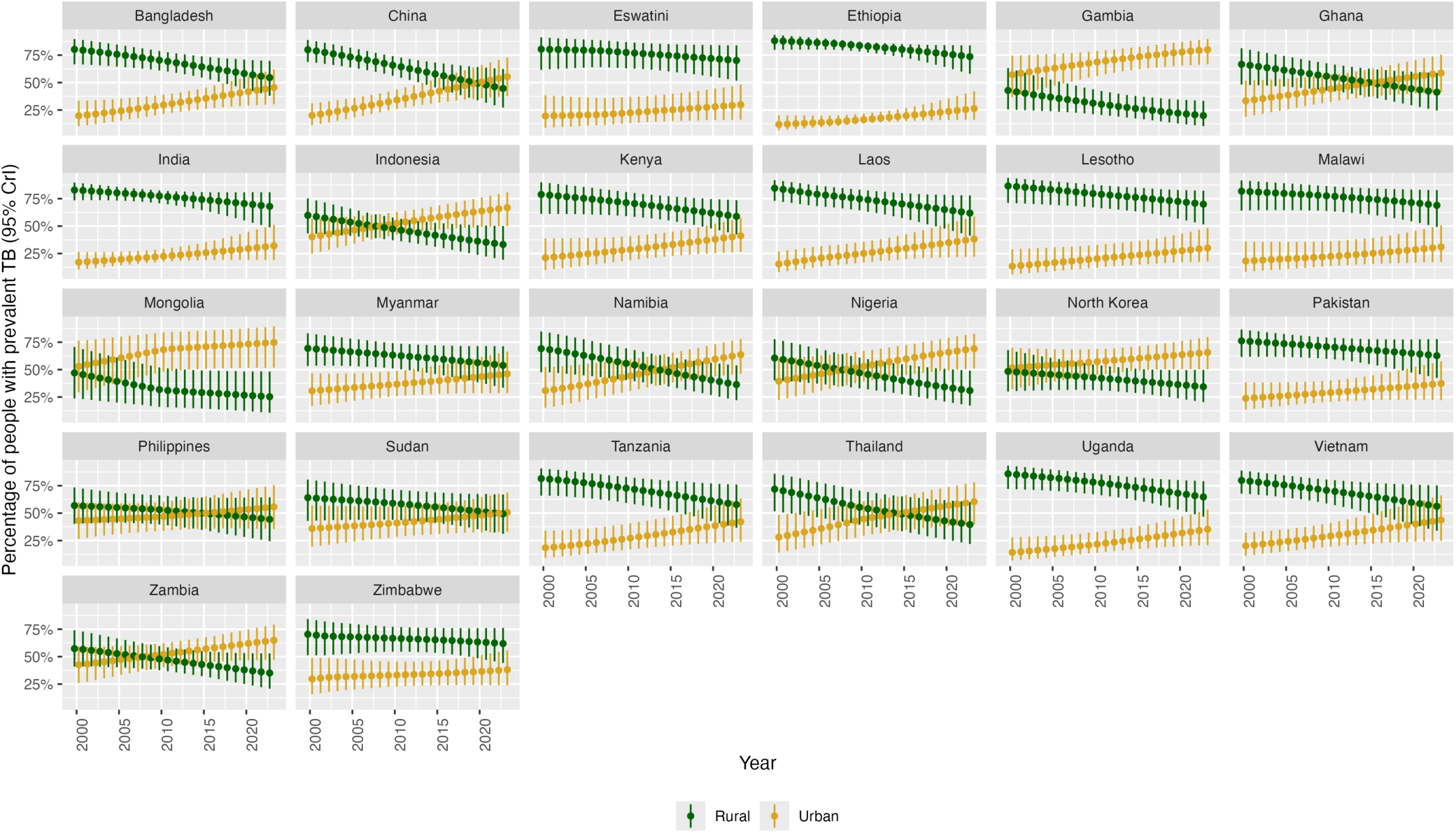
Percentage of people with prevalent TB in urban and rural areas in 26 countries: 2000-2023.

## Discussion

In this systematic review and meta-analysis of 46 national and subnational TB prevalence surveys from 26 low- and middle-income countries (together representing more than 4.6 billion people in 2023), we found that TB epidemics are becoming increasingly urbanised in both proportional and absolute terms, driven by rapidly urbanising demographic trends. Meta-analysis of smear-positive TB prevalence showed a faster rate of urbanisation than for bacteriologically-confirmed TB, emphasising the need to accelerate efforts to interrupt TB transmission within cities, as well as adapt to likely changing epidemiology in rural areas.

These findings highlight the importance of understanding local TB epidemiology to tailor public health strategies to context, which will be essential for accelerating progress toward the End TB Strategy targets and ensuring equitable health gains across all populations.

Urbanisation in low- and middle-income countries has been one of the major demographic shifts of the past century, with profound implications for the transmission and control of communicable diseases such as TB [71]. Between 2000 and 2023, the total populations of all study countries increased rapidly, with the proportion living in urban areas rising substantially in 25 of 26 countries, Zimbabwe being the exception.

For example, Uganda’s urban share grew by 81.1% (a 12 percentage-point increase, from 14.8 to 26.8%) and China’s by 80.0% (a 28.7 percentage-point increase from 35.9 to 64.6%). These shifts are largely driven by rural-to-urban migration, and in many high TB incidence countries this has resulted in substantial fractions of the urban population living in unplanned settlements and slums [71]. Rapid urban population growth, when combined with overcrowding, substandard housing, and under-resourced health systems, creates environments conducive to the transmission of infectious diseases, including TB [5,72].

Coinciding with these rapid demographic changes, we found that the urban-to-rural prevalence ratio of bacteriologically-confirmed TB likely increased over the 19 years during which surveys were conducted, with the rise being particularly pronounced for smear-positive disease. This trend cannot be explained by population redistribution alone: the ratio reflects a relative increase in prevalence, not simply a growing concentration of people in cities. In other words, TB burden within urban populations increased relative to rural populations, over and above demographic shifts toward urban living. However, this overall pattern masked substantial regional heterogeneity, with the direction and magnitude of the urban–rural gradient differing across Africa, South-East Asia, and the Western Pacific. The timing of transitions to predominantly urban TB epidemics also varied considerably between countries, reflecting differences in demographic change, health system capacity, and social conditions [72]. These findings suggest that demographic and epidemiological processes are acting in concert, with urban growth amplifying—but not solely determining, as emphasised by the absence of association with urbanisation indicators in meta-regression—the increasing concentration of TB in cities.

Regional patterns revealed substantial heterogeneity in the relative burden of TB between urban and rural populations. In the African Region, prevalence was higher in urban areas, with an overall urban-to-rural prevalence ratio exceeding one. In contrast, in the Western Pacific Region, prevalence was higher in rural populations, while in South-East Asia urban and rural prevalence was broadly similar. Despite higher rural prevalence in many Western Pacific countries, rapid urban population growth – particularly in China, Indonesia, and the Philippines – means that the absolute number and proportion of people with TB living in cities has become larger than in rural areas, and continues to rise. These shifts are compounded by rapid population ageing in the region [4,10], which disproportionately affects rural areas, as younger people migrate to cities for employment.

In countries in the African Region, the higher prevalence of TB in urban compared with rural populations is likely to reflect the combined influence of overlapping epidemics and demographic changes. Cities have been the epicentres of HIV epidemics in Africa [73,74], and HIV remains the strongest risk factor for progression from TB infection to disease [75]. The co-location of dense urban populations and concentrated HIV epidemics—along with other socioeconomic TB determinants—has therefore amplified TB transmission and disease incidence in cities. At the same time, many African countries are experiencing rapid population growth and urbanisation, with the most substantial increases occurring among younger age groups [76]. This demographic expansion fuels the absolute number of individuals at risk of TB, while high levels of mobility between rural and urban areas further complicate transmission dynamics and case detection [5].

In the South-East Asia Region, urban and rural TB prevalence appeared broadly similar, but this masks the enormous influence of India, which accounts for more than a quarter of the global TB burden [1]. India’s rapid urbanisation has produced vast, densely populated cities where TB transmission is sustained by poverty, overcrowding, and air pollution, yet large rural populations remain at risk due to limited access to timely diagnosis and care. Other countries in the region, including Indonesia, Myanmar, and Bangladesh, likely show similar dual challenges of intense transmission in urban centres and persistent barriers to care in rural areas. The absence of a strong urban–rural gradient in regional analyses therefore reflects both the scale of India’s contribution and the coexistence of high TB risks in different settings. For TB prevention and care in South-East Asia, addressing the needs of urban poor populations – particularly in informal settlements –while also strengthening rural primary care and social protection systems will be critical to reducing the overall burden.

Taken together, these dynamics suggest that TB control strategies must be informed by both demographic change and epidemiological context. In urban areas, interventions should prioritise the social determinants of TB—such as overcrowding, substandard housing, and poor air-quality [4] —while strengthening case-finding approaches, including targeted screening in high-risk groups [77], alongside systematic contact investigation and preventive therapy [78]. In high HIV-prevalence countries, particularly in Africa, public health efforts to combat TB in cities must integrate closely with HIV and other services [79] and adapt to the realities of youthful, highly mobile urban populations, with interventions spanning early diagnosis, preventive therapy, and targeted social support. In rural areas, where TB exposure may be more distal [80] and populations are ageing, strategies that improve access to routine screening within healthcare services, particularly for older adults, and that address risk factors for disease progression such as undernutrition [81], will likely be needed.

This study has several limitations. There is no unified global definition of urban and rural areas, and countries and regions often use varying criteria [72]. Only two studies were available from the Eastern Mediterranean Region (Pakistan and Sudan), each providing markedly different estimates of the urban-to-rural TB prevalence ratio; additional data from this, and other underrepresented WHO regions with high TB incidence countries (e.g. Region of the Americas, European), are needed. We excluded studies not published in English. To estimate numbers of people with prevalent TB and temporal trends, we relied on WHO incidence and case detection data combined with assumptions about disease duration. Although this approach has been widely used, including by WHO, transmission modelling methods may provide more robust estimates. We additionally projected estimates for numbers of people with prevalent TB beyond 2019 to 2023. All included prevalence surveys were conducted before 2019; estimates from the post-COVID-19 era are not available, and future prevalence surveys may be constrained by reductions in international funding [82].

In summary, TB epidemics are increasingly concentrated in urban populations both in proportional and absolute terms, likely driven by highly dynamic population changes over the past 20 years. In Africa, high urban TB prevalence, and proportions and numbers of people with prevalent TB in urban areas is likely to be amplified by rapid urbanisation, overlapping HIV epidemics, and rapid growth of young populations. In South-East Asia, the urban-to-rural prevalence ratio is moderated by the dominant contribution of the large rural population in India, and where both urban transmission and rural barriers to care shape the epidemic. In the Western Pacific, rural prevalence remains higher, yet rapid urban population growth and aging of the rural population, particularly in China, Indonesia and the Philippines, has rapidly increased the absolute number of urban people with prevalent TB. These findings highlight that both demographic change and local epidemiological context drive TB burden, underscoring the need for TB care and prevention strategies tailored to the specific social, demographic, and health system contexts of each setting.

## Data Availability

All data produced in the present work are contained in the manuscript

## Supporting information

**S1 Table:** Inclusion and exclusion eligibility criteria.

|  | Include | Exclude |
| --- | --- | --- |
| Population | Adults (≥ 15 years) or all-age samples from low- and middle-income countries (LMICs) as defined by the World Bank classification (2022-2023)<br>Studies allowing stratified analysis of active TB prevalence by sex (refer to this for full-text screening only) | Only includes symptomatic or healthcare- seeking individuals. Healthcare seeking can be for any disease.<br>Studies focused solely on occupational or university contexts.<br>Studies looking at latent TB prevalence.<br>Studies on congregate setting (occupational, prison, health facility, homeless shelter, etc.)<br>Research conducted in high-income countries.<br>Only children (< 15 years) |
| Comparator/<br>Context | TB disease prevalence<br>Total TB Prevalence: This includes all TB cases that are substantiated through bacteriological, radiological, or clinical evidence, capturing the full spectrum of the disease presentation.<br>Bacteriologically Confirmed TB: This subset is restricted to TB cases that have been confirmed through bacteriological methods such as smear microscopy, culture, or molecular diagnostics (e.g., Xpert MTB/RIF). Where the data allow, these cases will be further stratified into smear-positive and smear-negative.<br>TB Without Bacteriological Confirmation: This classification is reserved for TB cases diagnosed on the basis of radiological or clinical evidence in the absence of bacteriological confirmation. | Only extra-pulmonary TB (EPTB) due to the specialist nature of diagnosis.<br>Only DR-TB<br>Only latent TB infection prevalence |
| **Outcome** | Studies allowing stratified analysis of active TB prevalence by at least one of sex, urban/rural location, HIV status, and age group (refer to this for full-text screening only) | Unstratified active TB prevalence estimates. |
| **Study characteristics** | Prevalence surveys, whether stand-alone or part of larger studies (case-control, cohort studies, RCTs)<br>Studies with national to sub-national coverage | Routine notification or facility-based reporting<br>Contact tracing studies<br>Modelling studies |

**S2 Table:**
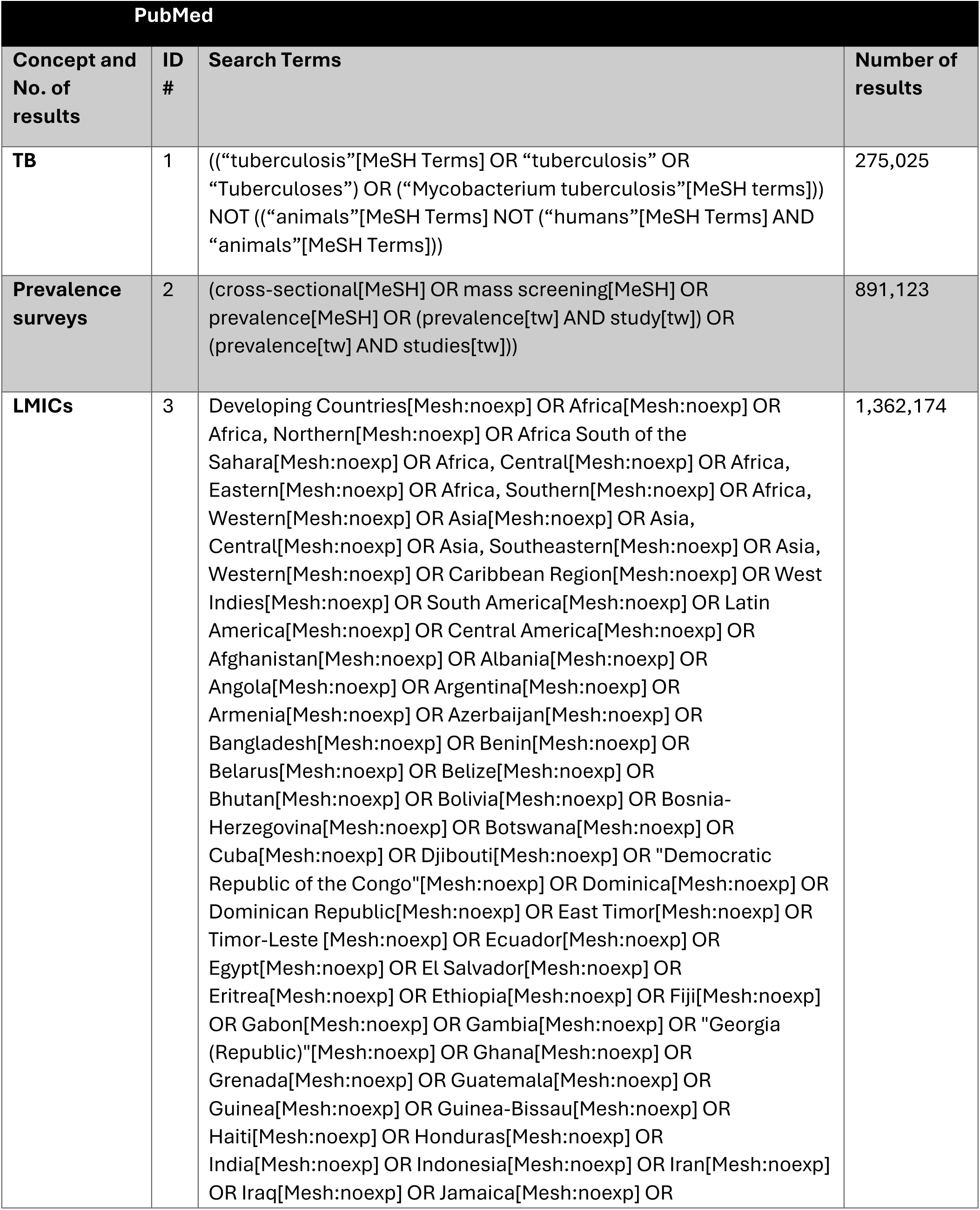
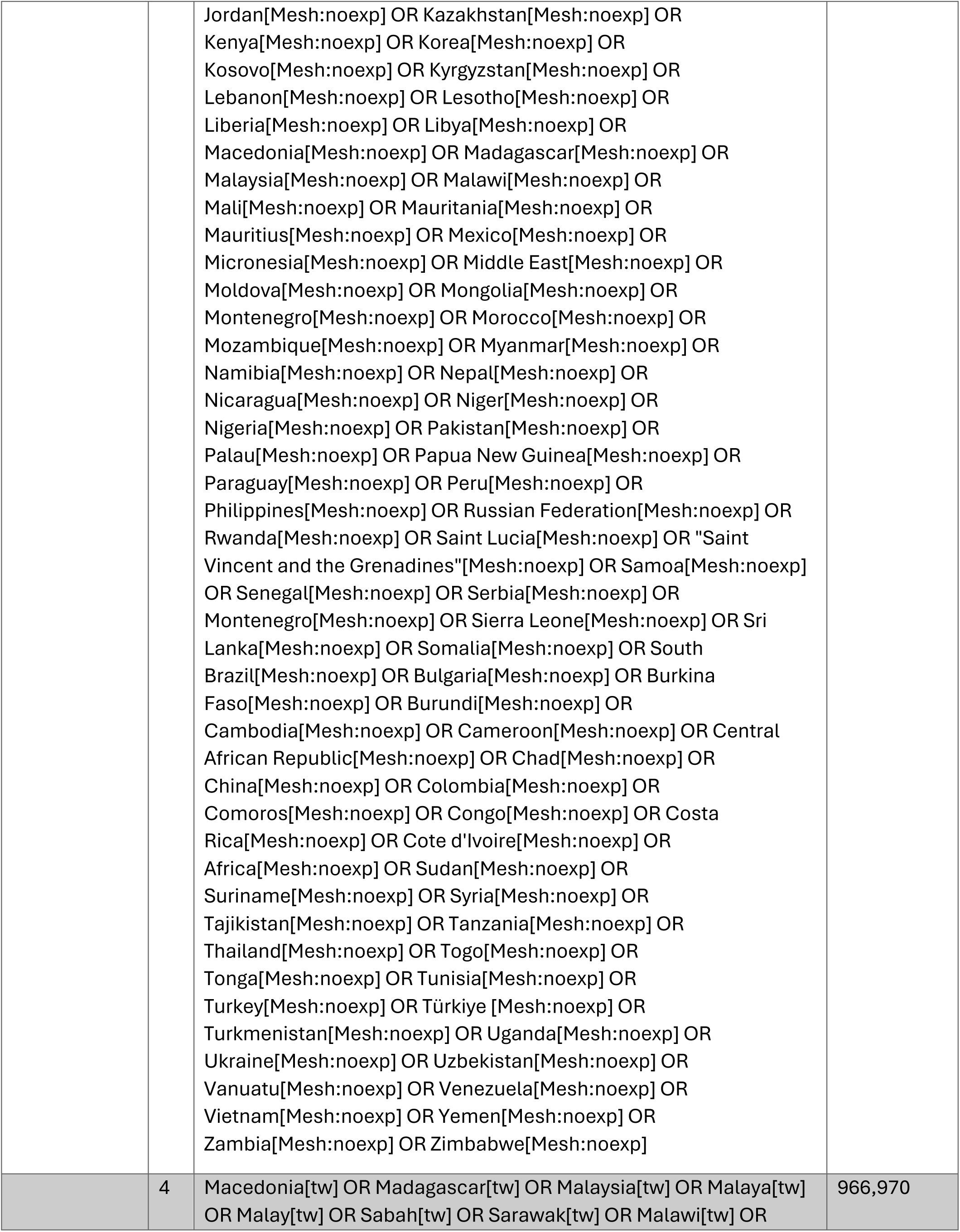
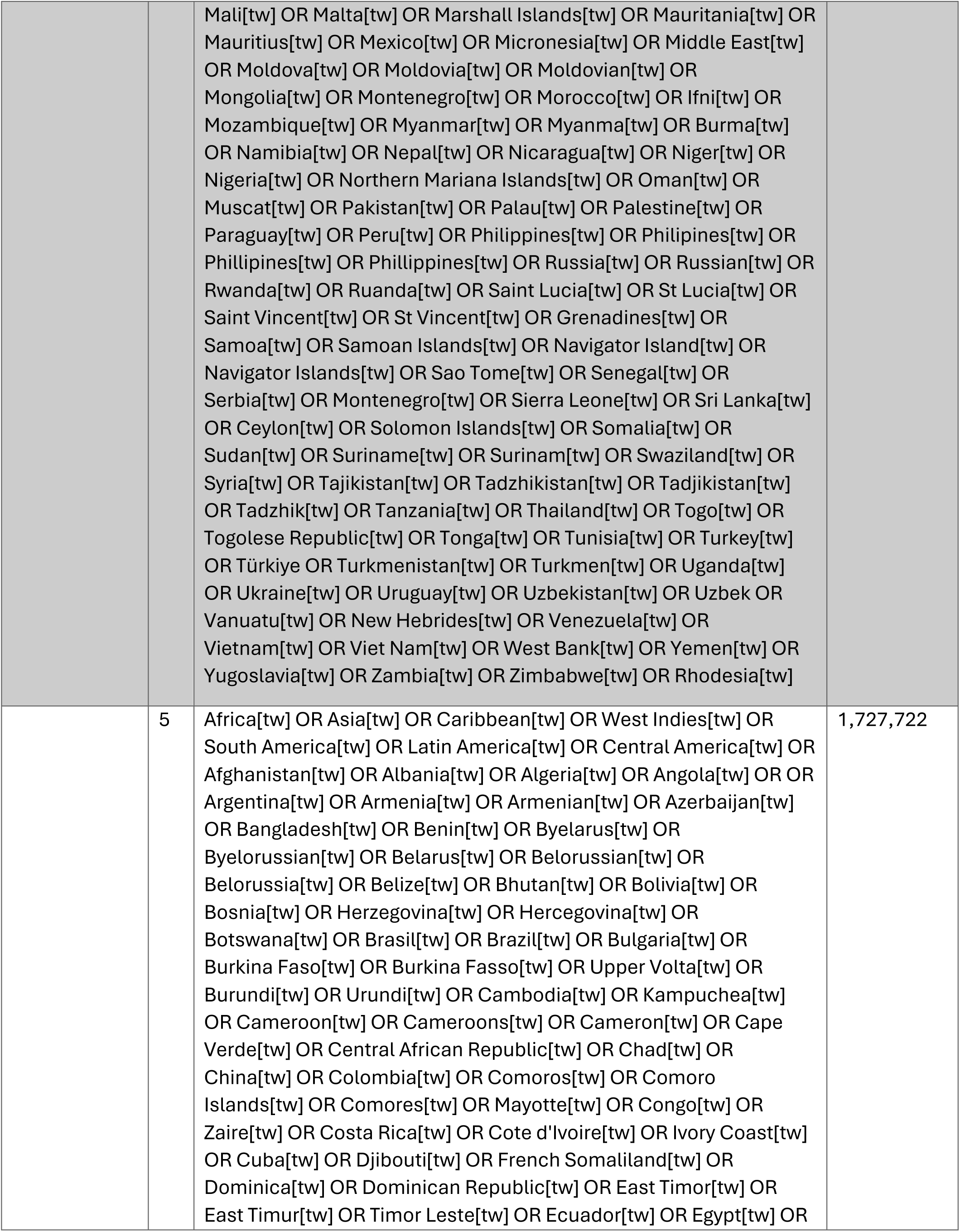
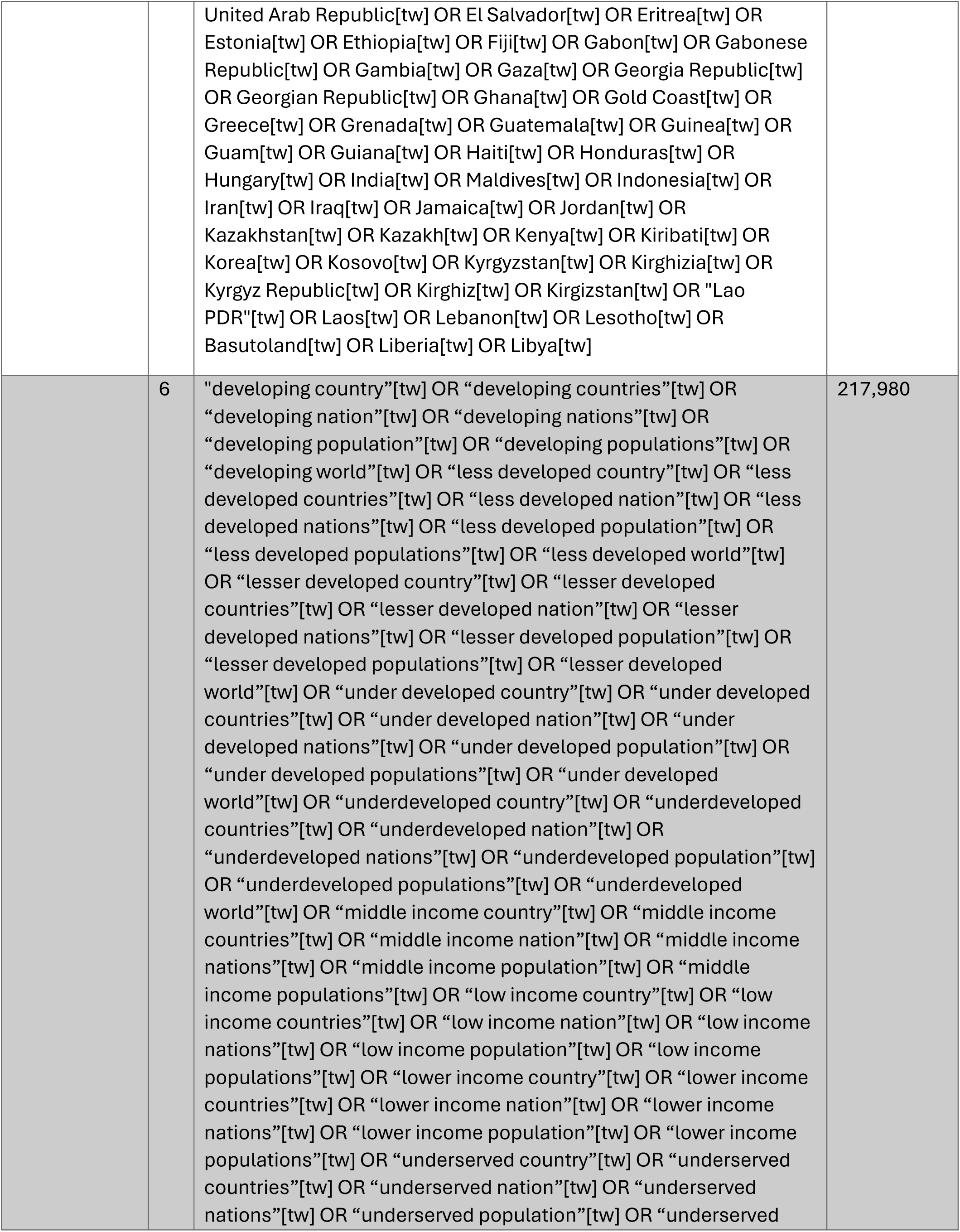
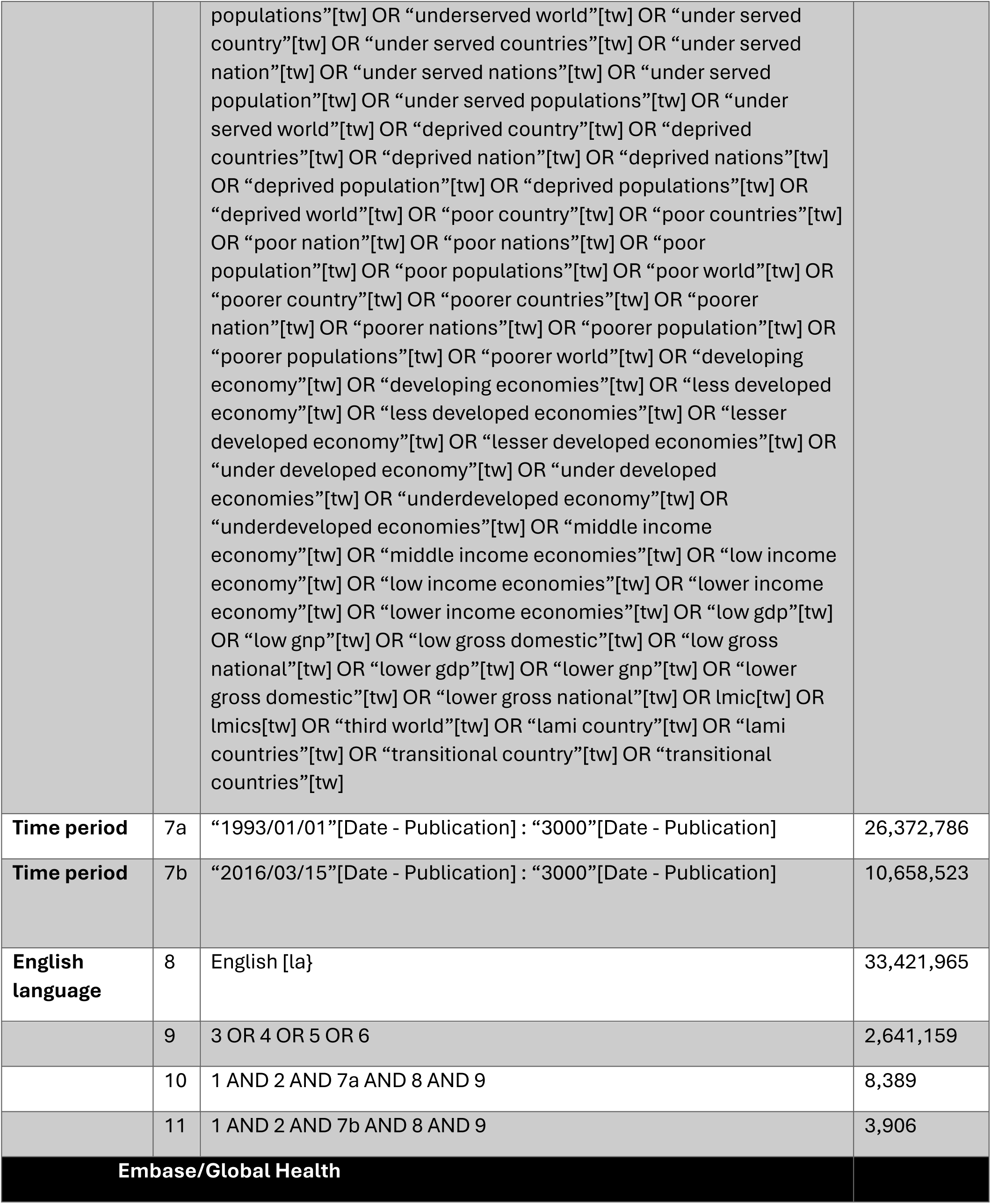
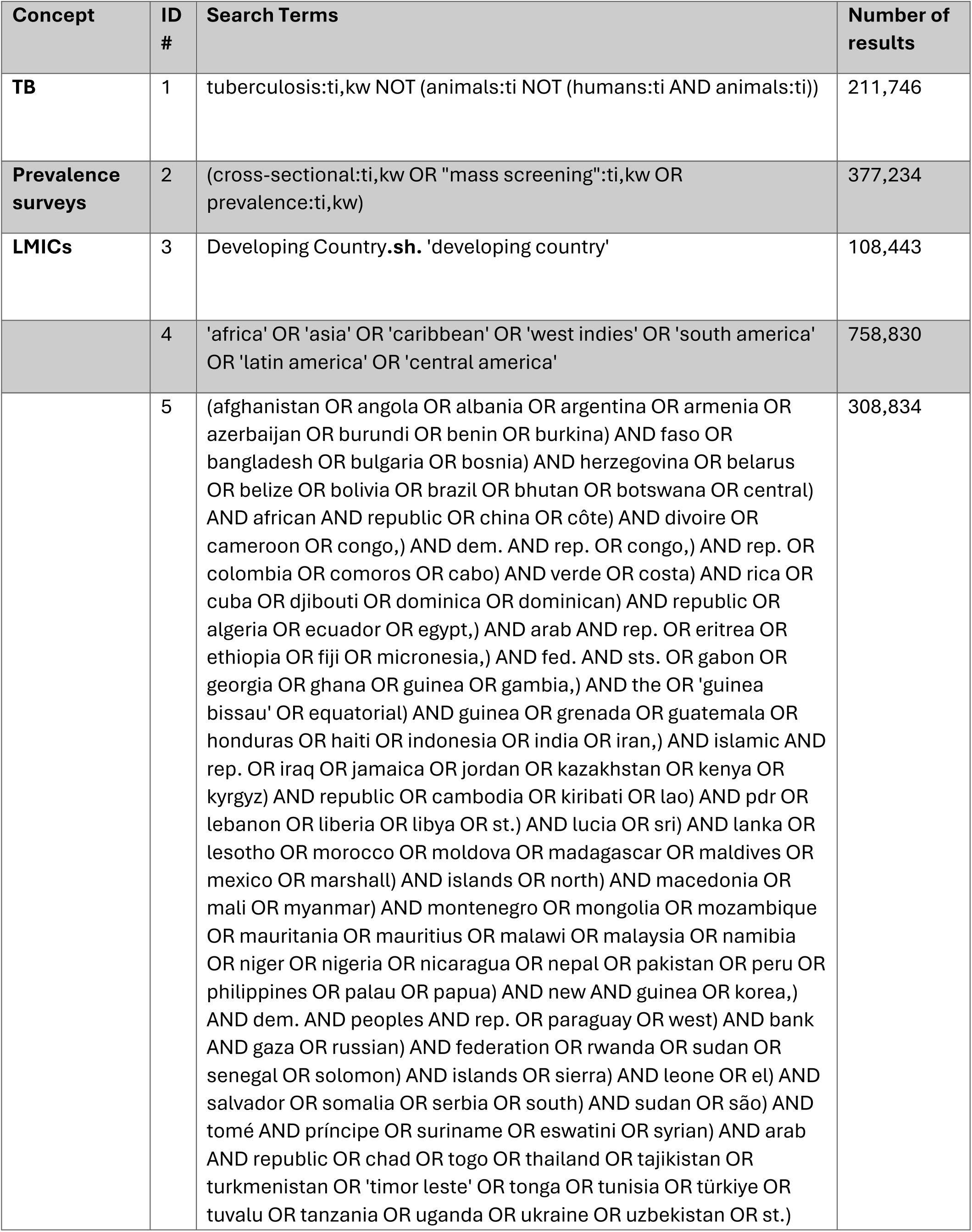
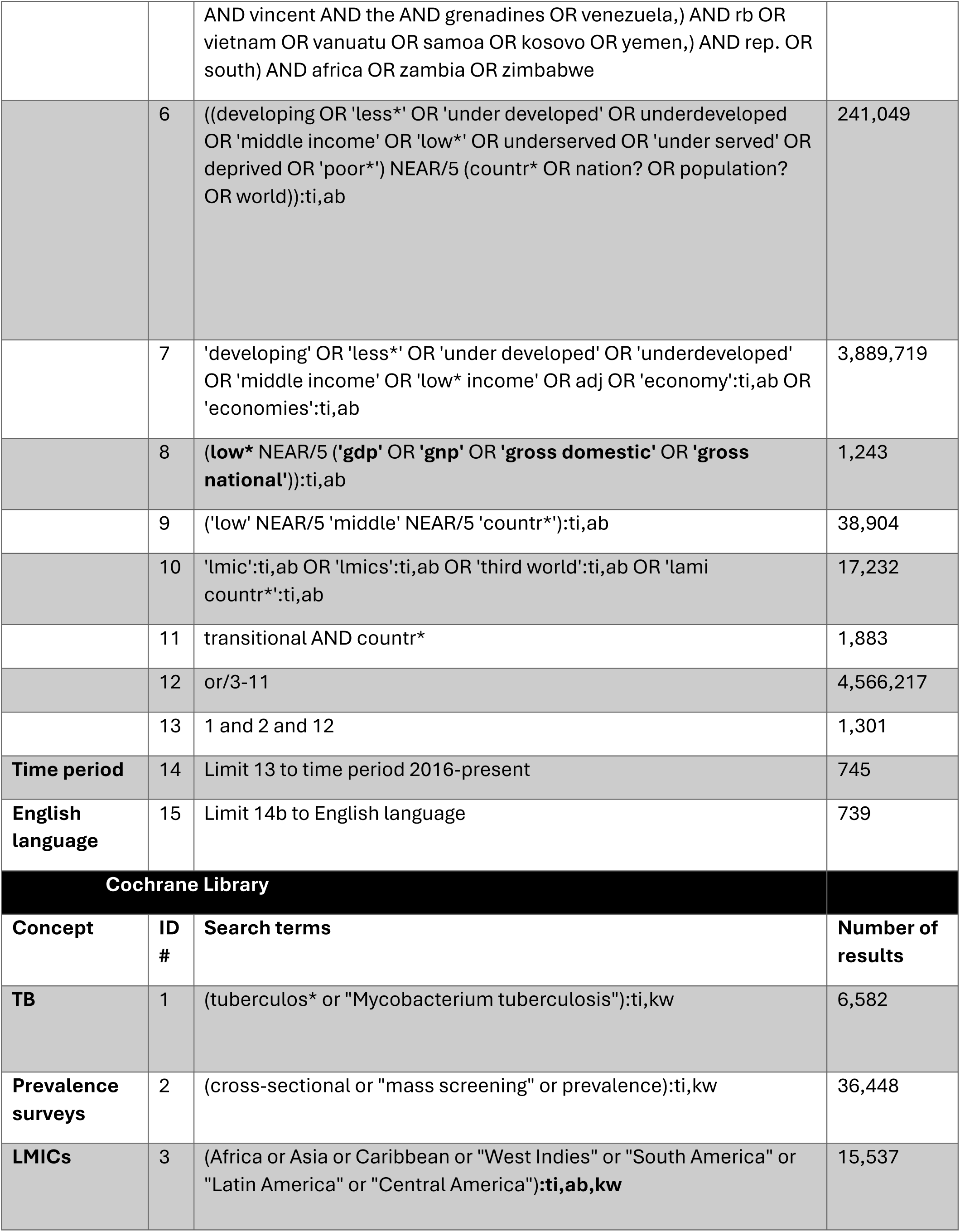
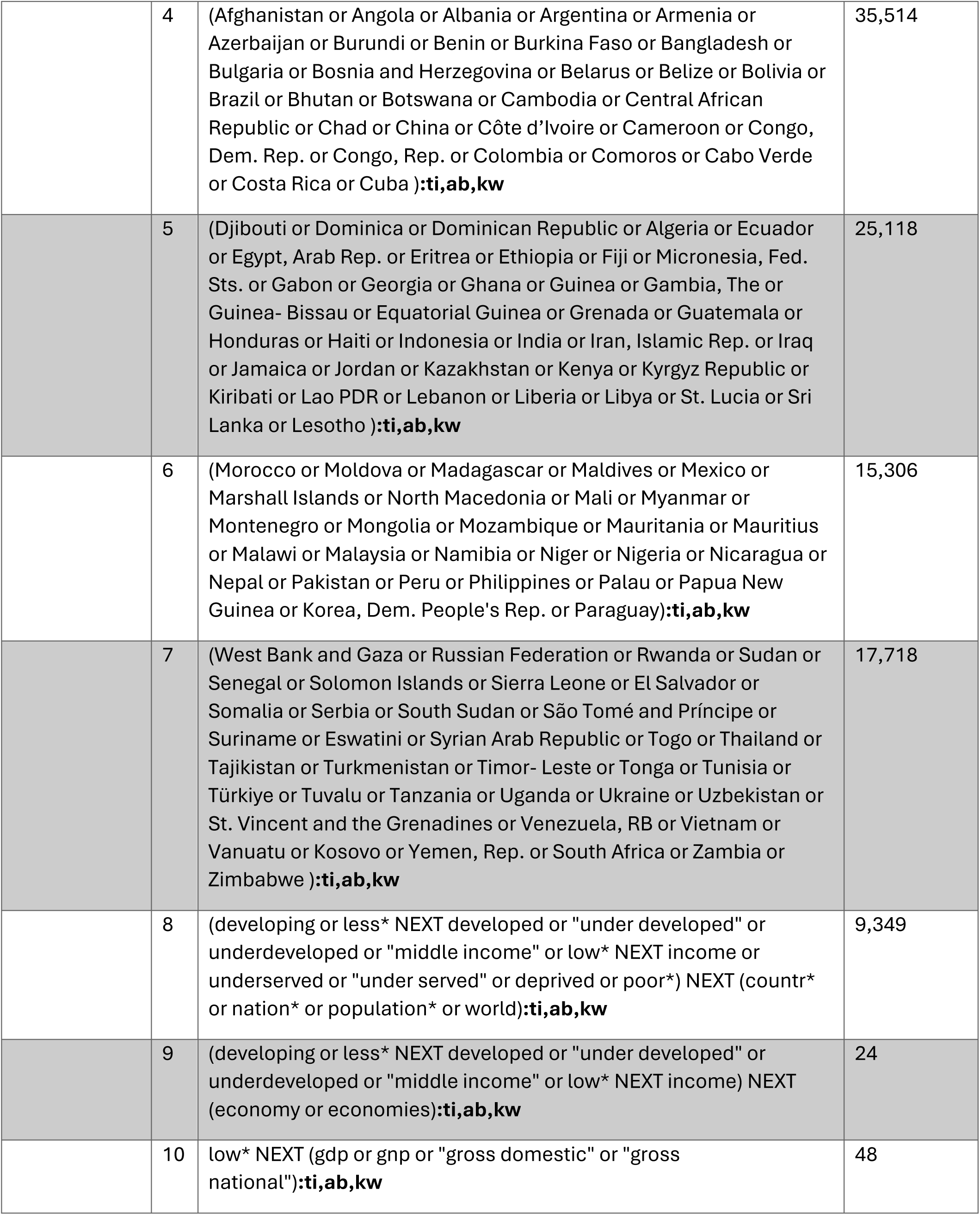
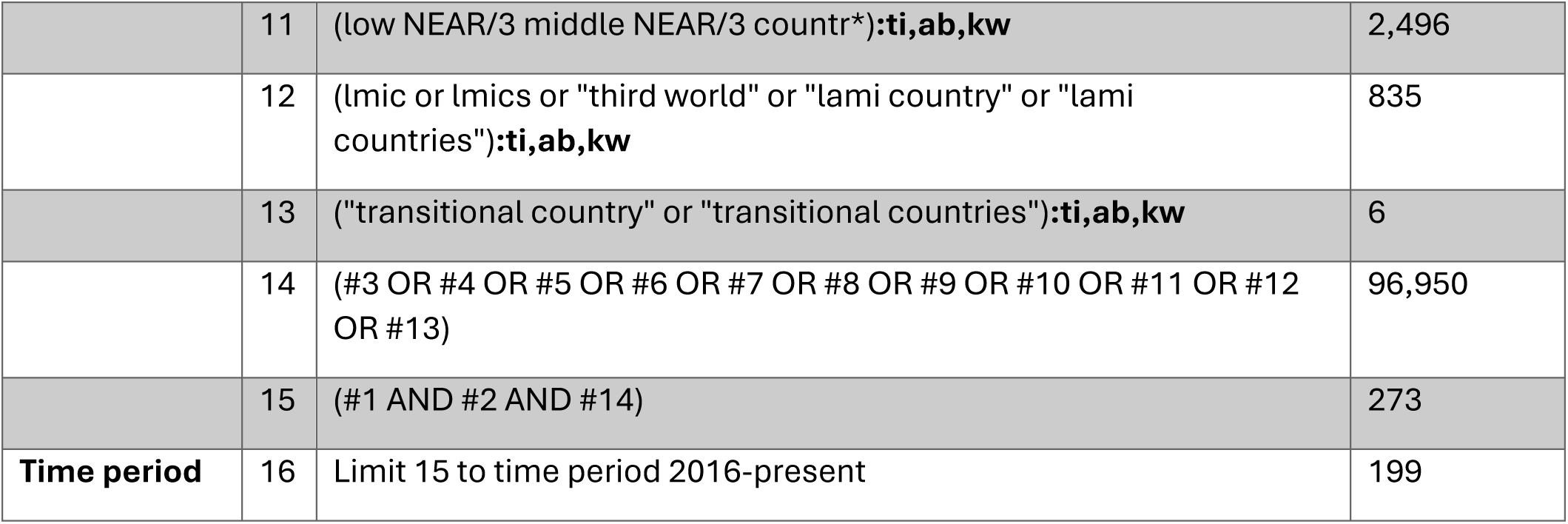
Search Strategy.

**S3 Fig.:**
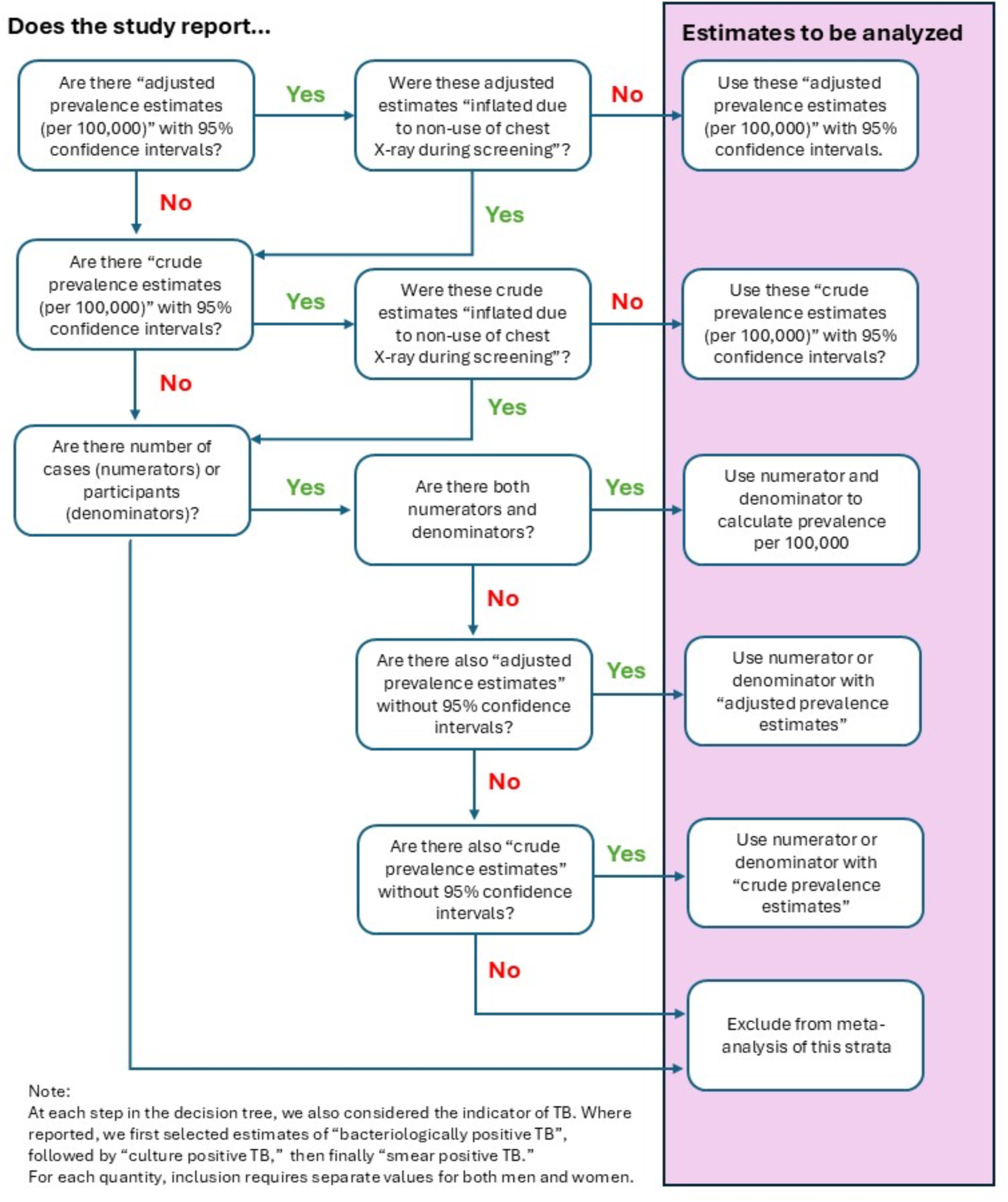
Hierarchy of TB prevalence survey estimates used in meta-analysis.

**S4 Text: Supplemental methods for estimating urban-to-rural prevalence ratio, and prevalence and burden of TB in urban and rural populations**

### Statistical modelling of urban-to-rural population ratio for burden estimation

To estimate the urban-to-rural TB prevalence burden (bacteriologically-confirmed), we fitted a Bayesian meta-analysis model to data extracted from 46 TB prevalence surveys from 26 countries conducted between 2000 and 2019. In models estimating global, and pooled regional prevalence ratios, we removed two countries from the Eastern Mediterranean Region with disparate estimates. For each survey, we calculated the log odds ratio of prevalence comparing urban to rural populations 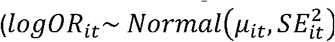 and its standard error (*SE_i_*_,*t*_), approximated from adjusted or un-adjusted 95% confidence intervals. For estimates without confidence intervals, we applied the normal approximation method of the binomial standard error.

We assumed log odds ratios were normally distributed around their expected values:

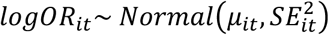

With the linear predictor modelled as:

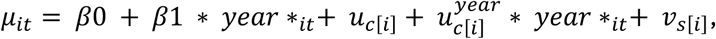

Where:

- *β*0 = overall intercept
- *β*1 = fixed effect of calendar year
- u_c[i]_ = random intercept for country c
- 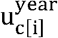 = random slope on year for country c
- *V_s_*_[*i*]_ = random intercept for study s

And priors weakly informative priors:

- *β*0 ∼ *Student* − *t*(7, 0, 1.5)
- *β*0 ∼ *Normal*(0, 10)
- *σ* ∼ *Exponential*(2)

Posterior summaries were based on means and 95% credible intervals. Study-, country- and year-specific urban-to-rural prevalence ratios were obtained by exponentiating posterior draws of *μ_it_*.

### Statistical modelling of incidence and case detection

To estimate the burden of prevalent TB in the 26 study countries, we obtained country-year estimates of TB incidence and case detection ratios from the WHO Global TB Database. For each country and year (2000– 2023), we extracted:

- The estimated incidence rate per 100,000 population (median and 95% uncertainty interval).
- The estimated case detection ratio (percentage of incident cases detected, 95% uncertainty interval).

From the World Bank and UN Population Prospects, we extracted mid-year national population denominators and distributions of urban and rural populations.

These data were merged to create country-year records of TB incidence, CDR, and population structure.

We modelled TB incidence and case detection jointly using a Bayesian multivariate regression model using brms as an interface to Stan. To allow for flexible temporal dynamics in incidence and case detection ratios, we used Gaussian process smooths for year by country.

For country *i* in year *t*:

### Incidence model

- Likelihood: 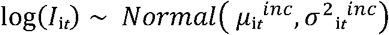
- Linear predictor: 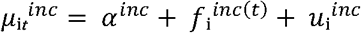

where *I*_i*t*_ is the WHO central incidence estimate, 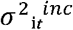 is derived from the incidence 95% uncertainty interval, 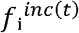 is a Gaussian process function of time with a country-specific kernel, and 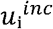 is a country random intercept.

### Case detection model

- Likelihood: 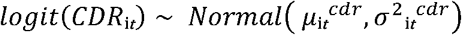
- Linear predictor: 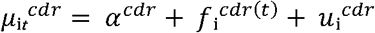

where *CDR*_i*t*_ is the WHO median case detection estimate, transformed to the logit scale, 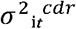 is derived from the 95% uncertainty interval, 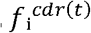 is a Gaussian process function of time with a country-specific kernel, and 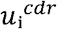 is a country random intercept.

### Priors

We used weakly informative priors, specifically:

- Intercept for the incidence model:

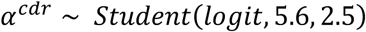

- Intercept for the CDR model:

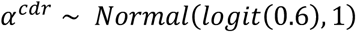

- Random effect standard deviations:

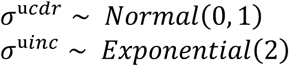

- Gaussian process hyperparameters (lengthscale, marginal variance) were given default inverse gamma and half-Student-t priors.

### Estimating prevalence from incidence and case detection

We converted posterior incidence and CDR predictions into prevalence estimates by applying disease duration assumptions.

Let:

- *d_T_* = mean duration of treated disease (years).
- *d_U_* = mean duration of untreated disease (years).

Then expected prevalence for country *i*, in year *t* is:

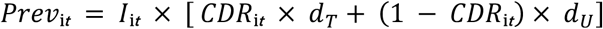

where *I*_i*t*_ is the estimated incidence rate per person-year.

- *d_T_* was fixed at 0.5 years. In sensitivity analyses, *d_T_* was sampled from a uniform distribution [0.25, 0.75].
- *d_U_* was drawn per posterior sample as a mixture of smear-positive TB (median 1.6 years, 95% CrI: 1.4–1.8) and smear-negative TB (median 5.4 years, 95% CrI: 3.4–8.2), with the mixing proportion sampled from a diffuse *Beta*(2,2) distribution.

### Urban and rural prevalence

We apportioned annual national prevalence estimates to urban and rural populations by multiplying by the proportion of the population in each country-year-rural/urban strata, and applying a posterior distribution of urban–rural prevalence ratios derived from meta-analysis of survey data.

Thus, for country *i*, year *t*:

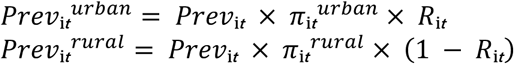

where *π*_i*t*_*^urban^* and *π*_i*t*_*^rural^* are the urban and rural population fractions, and *R*_i*t*_ is a posterior draw from the urban-to-rural prevalence ratio distribution.

### Model fitting

Models were fit in brms with 4 chains, each of 4000 iterations. Posterior predictive checks confirmed good model fit, with Gaussian process terms capturing non-linear temporal variation in both incidence and case detection across countries.

**S5 Table:** Summary characteristics of included studies.

| Country / Year | WHO Region | Case Definition | Diagnostic Tests | Urban n / N | Rural n / N | Urban Prevalence (per 100k) | Rural Prevalence (per 100k) | Crude Urban: Rural Ratio | Study Bias Assessment |
| --- | --- | --- | --- | --- | --- | --- | --- | --- | --- |
| Malawi (National) 2013-2014 | African Region | Adjusted Bacteriological TB Prevalence | Smear, Culture, Xpert MTB/RIF | 29/3654 | 103/27925 | 1,014.0 | 373.0 | 2.7 | Low |
| Ethiopia (Dabat District) 2010 | African Region | Count of Smear-Positive TB Cases | Smear | 6/5027 | 35/18563 | 119.4 | 188.5 | 0.6 | Moderate |
| Kenya (Karemo) NA | African Region | Count of Bacteriological TB Cases | Smear, Culture | 1/275 | 15/4729 | 363.6 | 317.2 | 1.1 | Moderate |
| The Gambia (National) 2012-2013 | African Region | Adjusted Bacteriological TB Prevalence | Smear, Culture | 37/17546 | 34/25554 | 266.0 | 109.0 | 2.4 | Low |
| Tanzania (National) 2011-2012 | African Region | Adjusted Bacteriological TB Prevalence | Smear, Culture, Xpert MTB/RIF | 17/6340 | 102/30889 | 279.0 | 315.0 | 0.9 | Moderate |
| Zambia (National) 2013-2014 | African Region | Adjusted Bacteriological TB Prevalence | Smear, Culture, Xpert MTB/RIF | NA*/16057 | NA/30042 | 993.0 | 460.0 | 2.2 | Low |
| Nigeria (National) 2012 | African Region | Adjusted Bacteriological TB Prevalence | Smear, Culture | NA/NA | NA/NA | 663.0 | 323.0 | 2.1 | Moderate |
| Ethiopia (National) 2010-2011 | African Region | Adjusted Bacteriological TB Prevalence | Smear, Culture | NA/7490 | NA/34952 | 273.0 | 273.0 | 1.0 | Low |
| Ethiopia (Tigray) 2011 | African Region | Adjusted Bacteriological TB Prevalence | Smear, Culture | 17/4624 | 13/7551 | 193.5 | 222.2 | 0.9 | Moderate |
| Zambia (Lusaka Province) 2005 | African Region | Adjusted Bacteriological TB Prevalence | Smear, Culture | 51/NA | 28/NA | 1,200.0 | 650.0 | 1.8 | Moderate |
| Uganda (National) 2014-2015 | African Region | Adjusted Bacteriological TB Prevalence | Smear, Culture, Xpert MTB/RIF | 80/17338 | 80/23816 | 504.0 | 370.0 | 1.4 | Low |
| Namibia (National) 2017-2018 | African Region | Adjusted Bacteriological TB Prevalence | Culture, Xpert MTB/RIF | 54/12178 | 65/17317 | 537.3 | 391.4 | 1.4 | Low |
| Eswatini (National) 2018-2019 | African Region | Adjusted Bacteriological TB Prevalence | Culture, Xpert MTB/RIF | NA/1196 | NA/8742 | 385.0 | 344.0 | 1.1 | Low |
| Lesotho (National) 2019 | African Region | Adjusted Bacteriological TB Prevalence | Culture, Xpert Ultra | 40/8230 | 85/12490 | 453.0 | 670.0 | 0.7 | Low |
| Kenya (National) 2015-2016 | African Region | Adjusted Bacteriological TB Prevalence | Smear, Culture, Xpert MTB/RIF | NA/19444 | NA/43606 | 760.0 | 453.0 | 1.7 | Low |
| Zimbabwe (National) 2014 | African Region | Adjusted Bacteriological TB Prevalence | Smear, Culture, Xpert MTB/RIF | 31/9097 | 76/24639 | 355.0 | 337.0 | 1.1 | Low |
| Ethiopia (Hetosa District) 2013 | African Region | Adjusted Bacteriological TB Prevalence | Smear, Culture | 9/5903 | 36/27170 | 153.0 | 125.0 | 1.2 | Low |
| Ghana (National) 2013 | African Region | Adjusted Bacteriological TB Prevalence | Smear, Culture, Xpert MTB/RIF | NA/33122 | NA/28604 | 293.0 | 429.0 | 0.7 | Low |
| The Gambia (National) 2011-2013 | African Region | Adjusted Bacteriological TB Prevalence | Smear, Culture | NA/17546 | NA/25554 | 266.0 | 109.0 | 2.4 | Low |
| Sudan (National) 2013-2014 | Eastern Mediterranean Region | Adjusted Bacteriological TB Prevalence | Smear, Culture | NA/29912 | NA/53290 | 275.6 | 136.0 | 2.0 | Low |
| Pakistan (National) 2010-2011 | Eastern Mediterranean Region | Adjusted Bacteriological TB Prevalence | Smear, Culture, Xpert MTB/RIF | 117/47809 | 224/58104 | 310.0 | 470.5 | 0.7 | Moderate |
| India (Kanpur Nagar District) 2008-2010 | South-East Asian Region | Adjusted Bacteriological TB Prevalence | Smear, Culture | NA/NA | NA/NA | 328.8 | 810.9 | 0.4 | Low |
| India (Gujarat State) 2011-2012 | South-East Asian Region | Adjusted Bacteriological TB Prevalence | Smear, Culture | 102/32940 | 156/39068 | 389.0 | 487.3 | 0.8 | Low |
| India (Wardha District) 2007-2009 | South-East Asian Region | Adjusted Bacteriological TB Prevalence | Smear, Culture | 19/14030 | 67/36302 | 153.8 | 189.1 | 0.8 | Low |
| Thailand (National) 2012 | South-East Asian Region | Unadjusted Bacteriological TB Prevalence | Smear, Culture | 21/NA | 121/NA | 240.5 | 224.9 | 1.1 | NA |
| Bangladesh (National) 2007-2009 | South-East Asian Region | Adjusted Smear-Positive TB Prevalence | Smear, Culture | 13/26046 | 20/26052 | 51.1 | 86.0 | 0.6 | Moderate |
| Indonesia (National) 2004 | South-East Asian Region | Adjusted Smear-Positive TB Prevalence | Smear, Culture | 33/23150 | 47/27004 | 102.0 | 106.0 | 1.0 | Moderate |
| India (Faridabad District) 2008-2009 | South-East Asian Region | Count of Bacteriological TB Cases | Smear, Culture | 40/57338 | 60/41261 | 69.8 | 145.4 | 0.5 | Low |
| India (Jabalpur District) 2009-2010 | South-East Asian Region | Unadjusted Smear-Positive TB Prevalence | Smear, Culture | NA/NA | NA/NA | 153.9 | 348.9 | 0.4 | Low |
| Myanmar (National) 2009-2010 | South-East Asian Region | Adjusted Bacteriological TB Prevalence | Smear, Culture | 103/11254 | 208/40113 | 903.2 | 526.8 | 1.7 | Low |
| India (Mohali District) 2008-2010 | South-East Asian Region | Unadjusted Bacteriological TB Prevalence | Smear, Culture | NA/34161 | NA/51609 | 11.7 | 32.9 | 0.4 | Low |
| Myanmar (National) 2017-2018 | South-East Asian Region | Count of Bacteriological TB Cases | Smear, Culture, Xpert Ultra | 136/24137 | 186/42343 | 563.5 | 439.3 | 1.3 | Moderate |
| DPR Korea (National) 2015-2016 | South-East Asian Region | Adjusted Bacteriological TB Prevalence | Smear, Culture | 194/34413 | 143/23123 | 577.0 | 659.0 | 0.9 | Low |
| Bangladesh (National) 2015-2016 | South-East Asian Region | Adjusted Bacteriological TB Prevalence | Smear, Culture, Xpert MTB/RIF | 107/34814 | 171/63896 | 316.0 | 270.0 | 1.2 | Low |
| Indonesia (National) 2013-2014 | South-East Asian Region | Adjusted Bacteriological TB Prevalence | Smear, Culture, Xpert MTB/RIF | 228/31871 | 198/36073 | 845.8 | 674.2 | 1.3 | Low |
| China (National) 2000 | Western Pacific Region | Adjusted Bacteriological TB Prevalence | Smear, Culture | NA/NA | NA/NA | 152.0 | 225.0 | 0.7 | Low |
| Viet Nam (National) 2006-2007 | Western Pacific Region | Adjusted Bacteriological TB Prevalence | Smear, Culture | 72/26357 | 197/67822 | 282.0 | 344.0 | 0.8 | Low |
| China (Shandong Province) 2010 | Western Pacific Region | Adjusted Bacteriological TB Prevalence | Smear, Culture | NA/26101 | NA/28178 | 9.0 | 43.1 | 0.2 | Low |
| China (National) 2010 | Western Pacific Region | Adjusted Bacteriological TB Prevalence | Smear, Culture | NA/NA | NA/NA | 73.0 | 163.0 | 0.4 | Low |
| Philippines (National) 2007 | Western Pacific Region | Adjusted Bacteriological TB Prevalence | Smear, Culture | NA/8331 | NA/11635 | 670.0 | 650.0 | 1.0 | Moderate |
| Lao Pdr (National) 2010-2011 | Western Pacific Region | Adjusted Bacteriological TB Prevalence | Smear, Culture | 42/10209 | 195/29003 | 436.0 | 663.0 | 0.7 | Low |
| Philippines (National) 2016 | Western Pacific Region | Count of Bacteriological TB Cases | Smear, Culture, Xpert MTB/RIF | 244/21526 | 222/25163 | 1,133.5 | 882.2 | 1.3 | Low |
| Mongolia (National) 2014-2015 | Western Pacific Region | Count of Bacteriological TB Cases | Smear, Culture, Xpert MTB/RIF | 142/27112 | 106/23197 | 523.8 | 457.0 | 1.1 | Low |
| China (Multiple Areas) 2013 | Western Pacific Region | Unadjusted Bacteriological TB Prevalence | Smear, Culture | 14/12933 | 48/21336 | 108.2 | 225.0 | 0.5 | Moderate |
| Viet Nam (National) 2017-2018 | Western Pacific Region | Count of Bacteriological TB Cases | Smear, Culture, Xpert MTB/RIF | 90/18656 | 131/43107 | 482.4 | 303.9 | 1.6 | Low |

| Country /<br>Year | WHO<br>Region | Case<br>Definition | Diagnostic<br>Tests | Urban n /<br>N | Rural n / N | Urban<br>Prevalence<br>(per<br>100k) | Rural<br>Prevalence<br>(per<br>100k) | Crude<br>Urban:<br>Rural<br>Ratio | Study<br>Bias<br>Assessment |
| --- | --- | --- | --- | --- | --- | --- | --- | --- | --- |
| China<br>(Xinjiang<br>Uyghur<br>Region)<br>2010-2011 | Western<br>Pacific<br>Region | Adjusted<br>Bacteriological<br>TB<br>Prevalence | Smear,<br>Culture | NA/8405 | NA/12104 | 109.0 | 506.0 | 0.2 | Low |

\*NA: not reported

**S6 Table:**
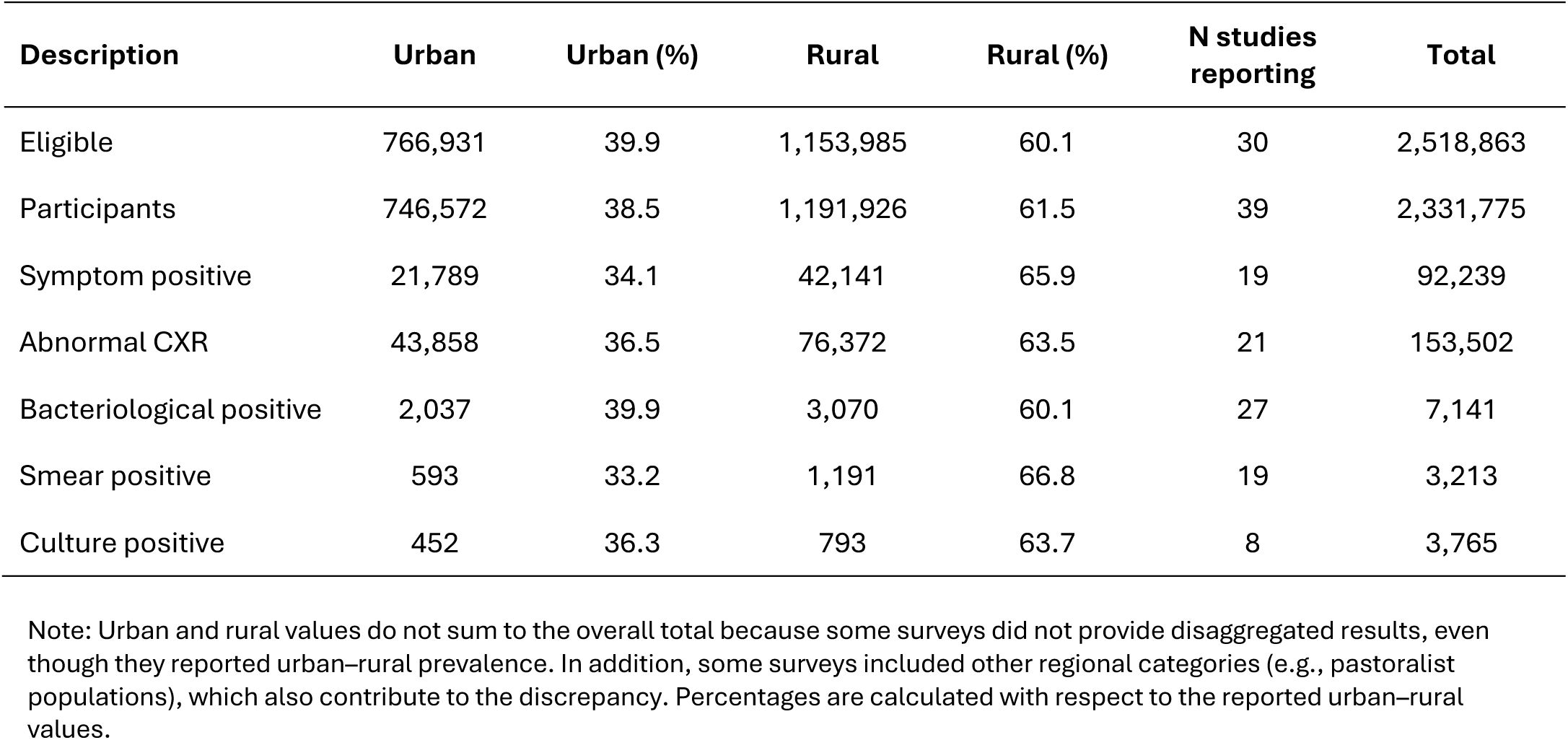
Descriptive Summary of Included Studies.

**S7 Figure:**
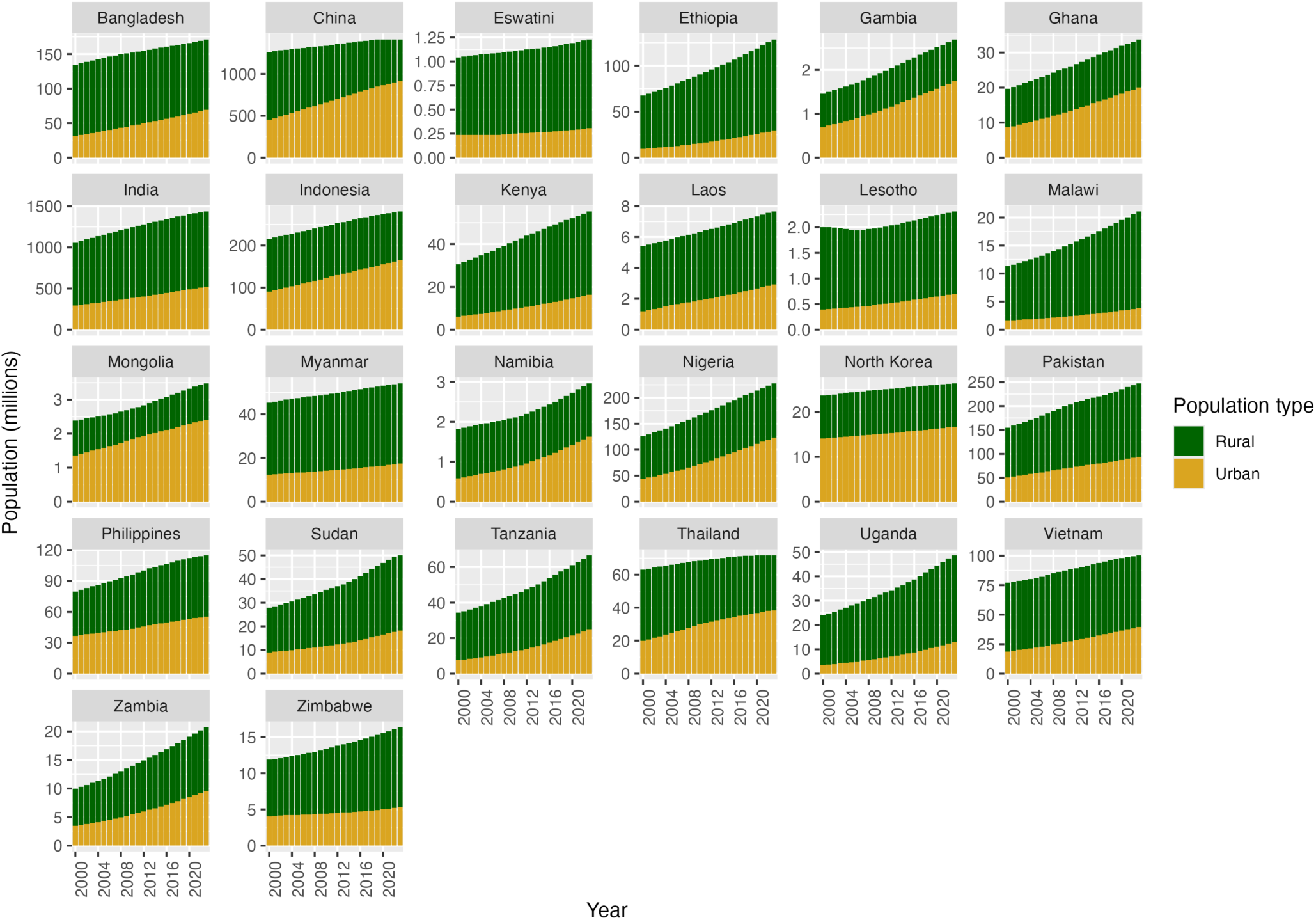
Urban and rural populations in 26 study countries between 2000 and 2023.

**S8 Figure:**
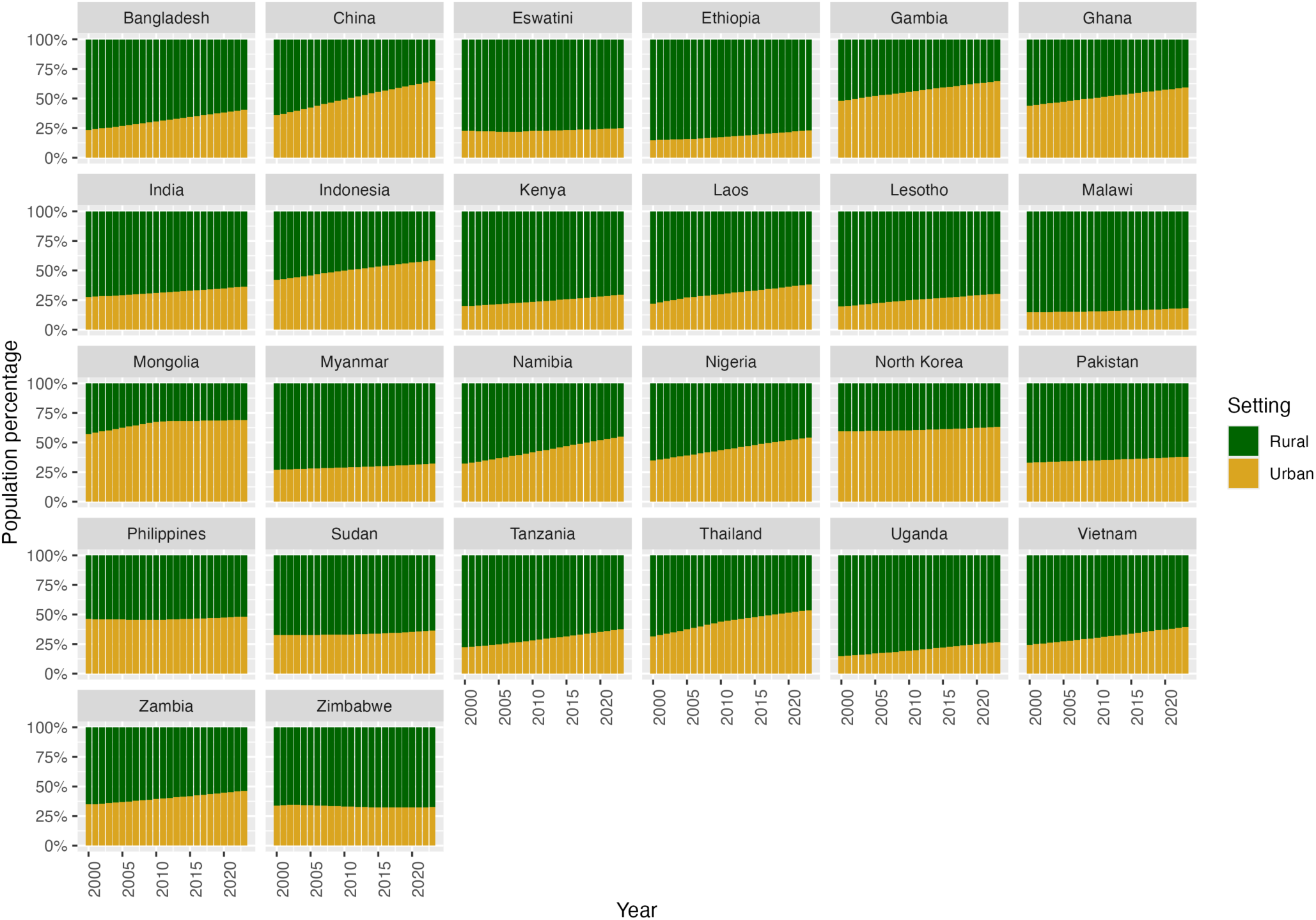
Urban and rural populations percentages in 26 study countries between 2000 and 2023.

**S9 Figure:**
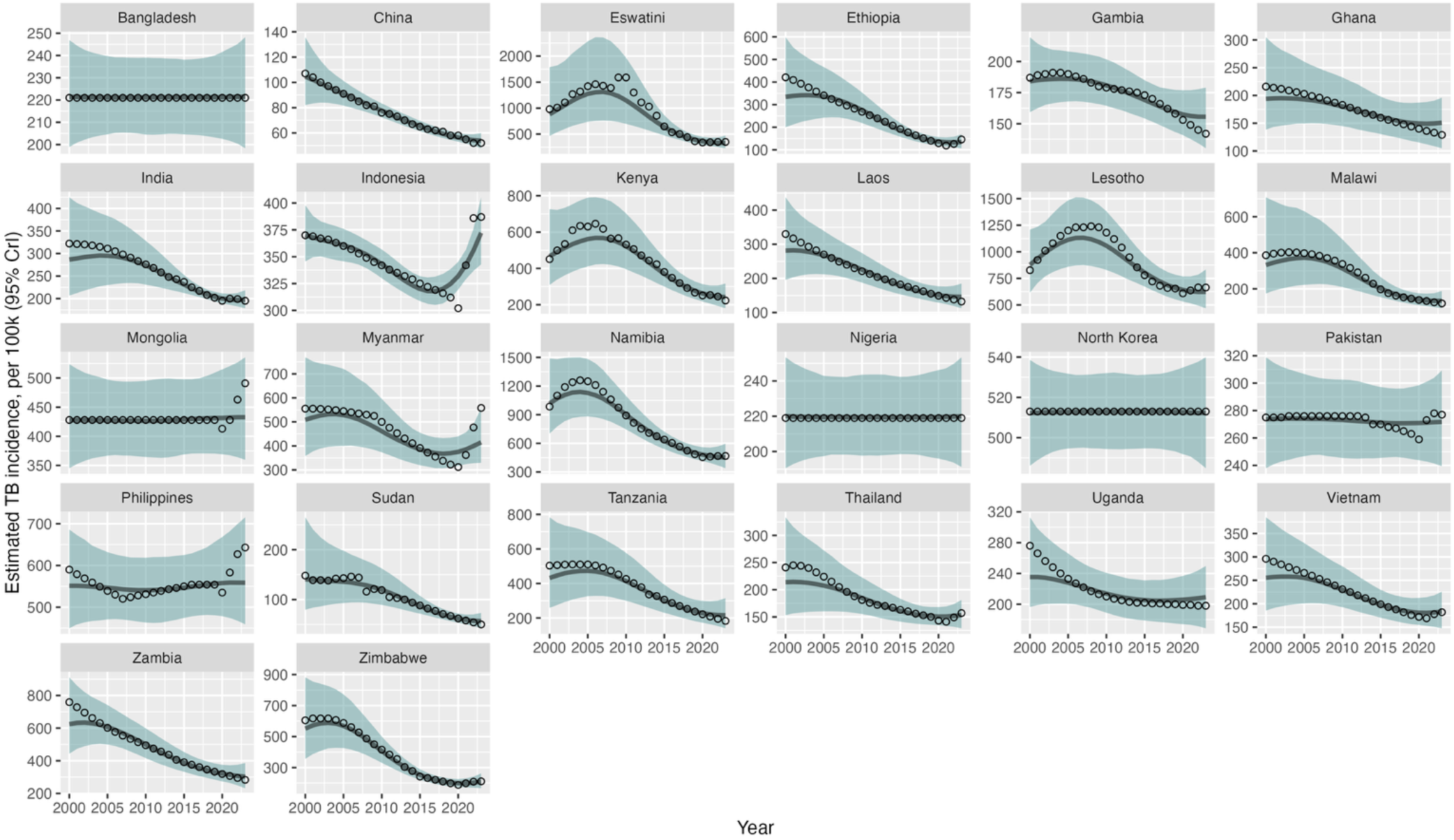
Estimated TB incidence in 26 study countries between 2000 and 2023. Black circles are central estimates, reported by WHO; blue line and bands are estimated from a Bayesian multivariate regression model of incidence and case detection ratio data.

**S10 Figure:**
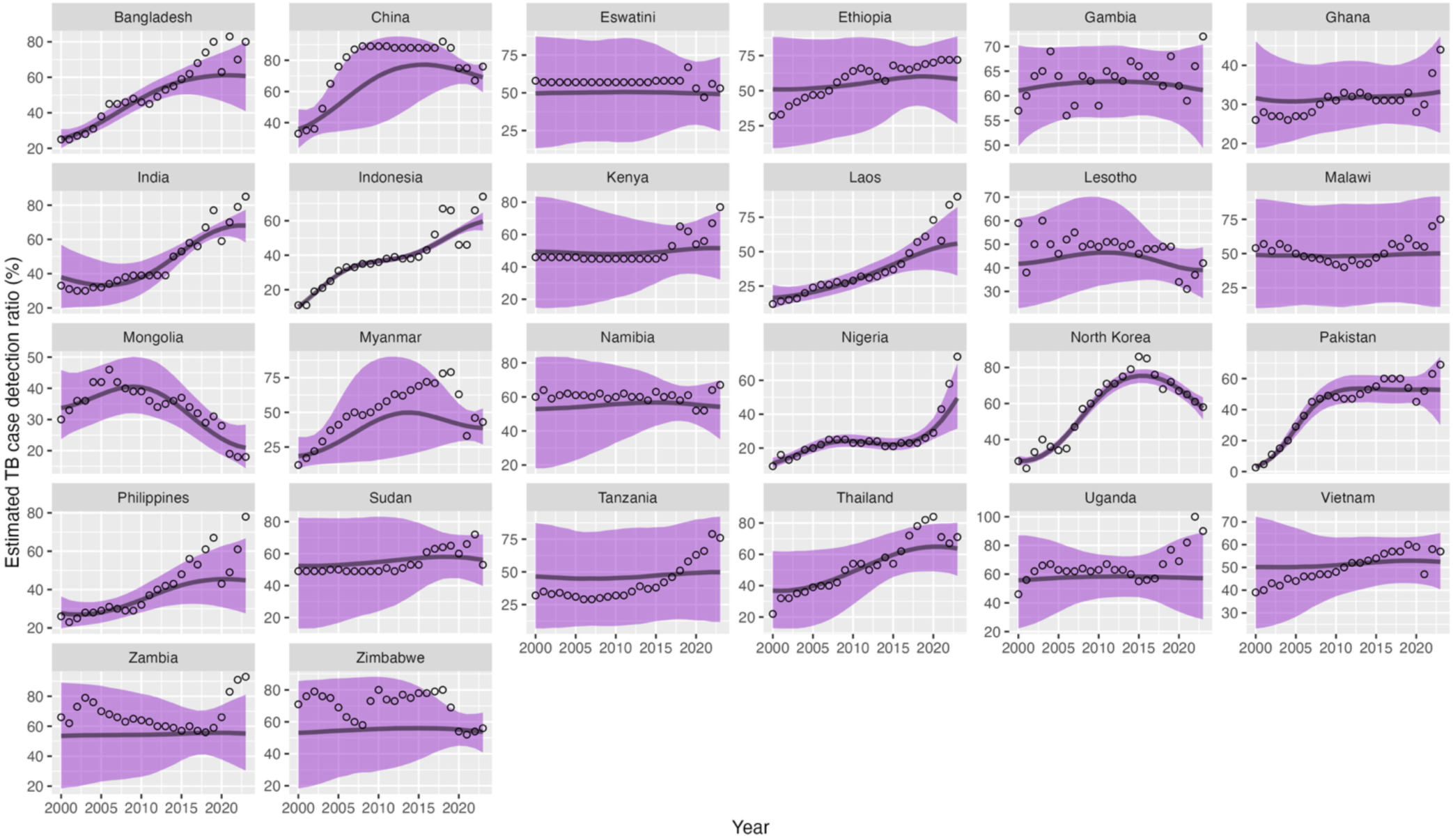
Estimated TB case detection ratio in 26 study countries between 2000 and 2023. Black circles are central estimates, reported by WHO; purple line and bands are estimated from a Bayesian multivariate regression model of incidence and case detection ratio data.

**S11 Figure:**
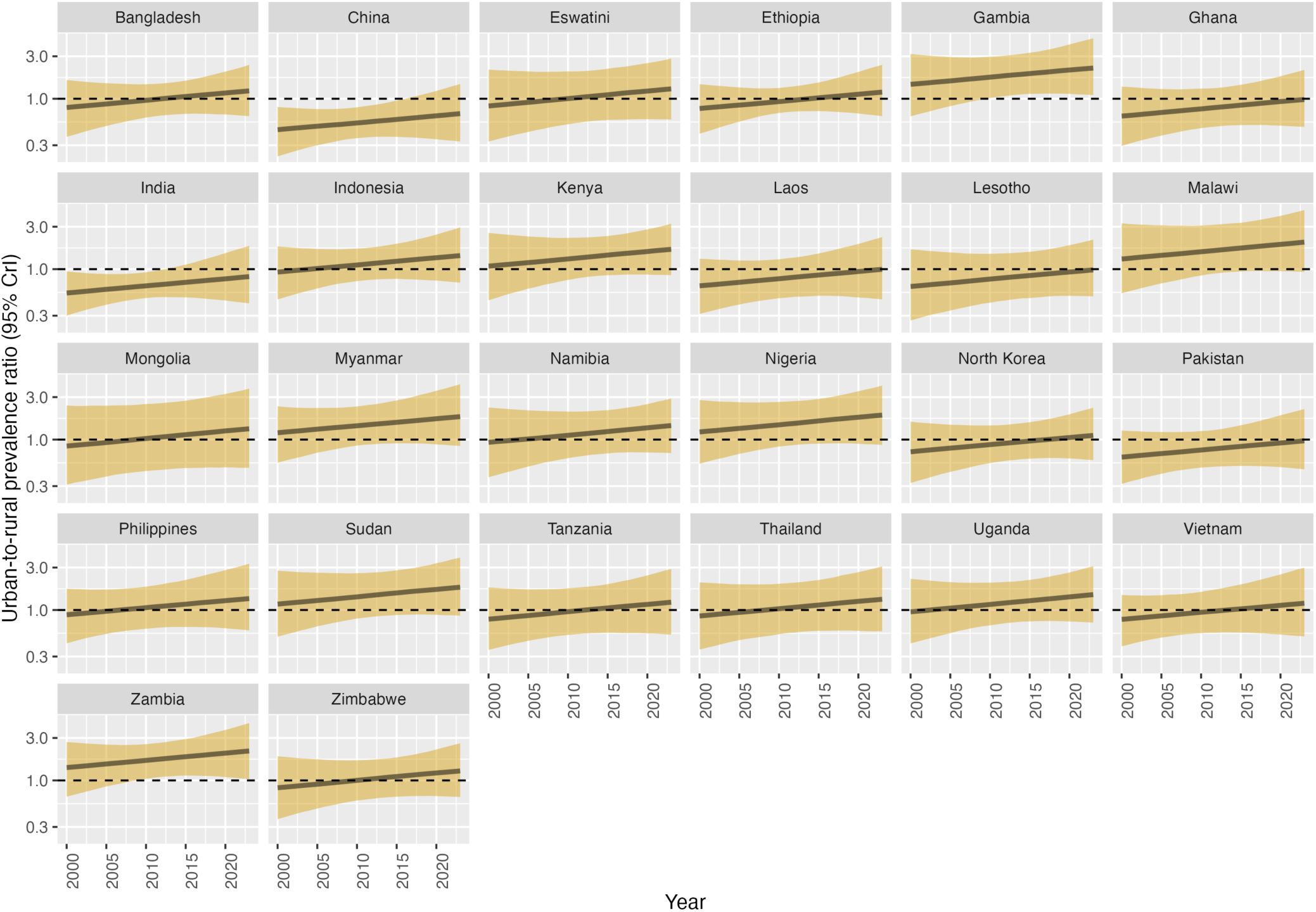
Predicted urban-to-rural prevalence ratio in bacteriologically-confirmed TB in 26 study countries: 2000-2023.

**S12 Table:**
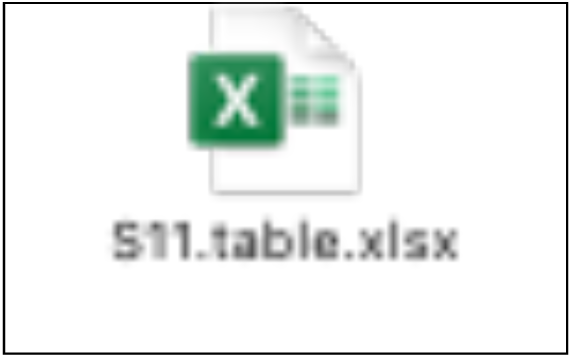
Estimated numbers and percentage of prevalent TB in urban and rural areas in 26 study countries between 2000 and 2023.

**S13 Figure:**
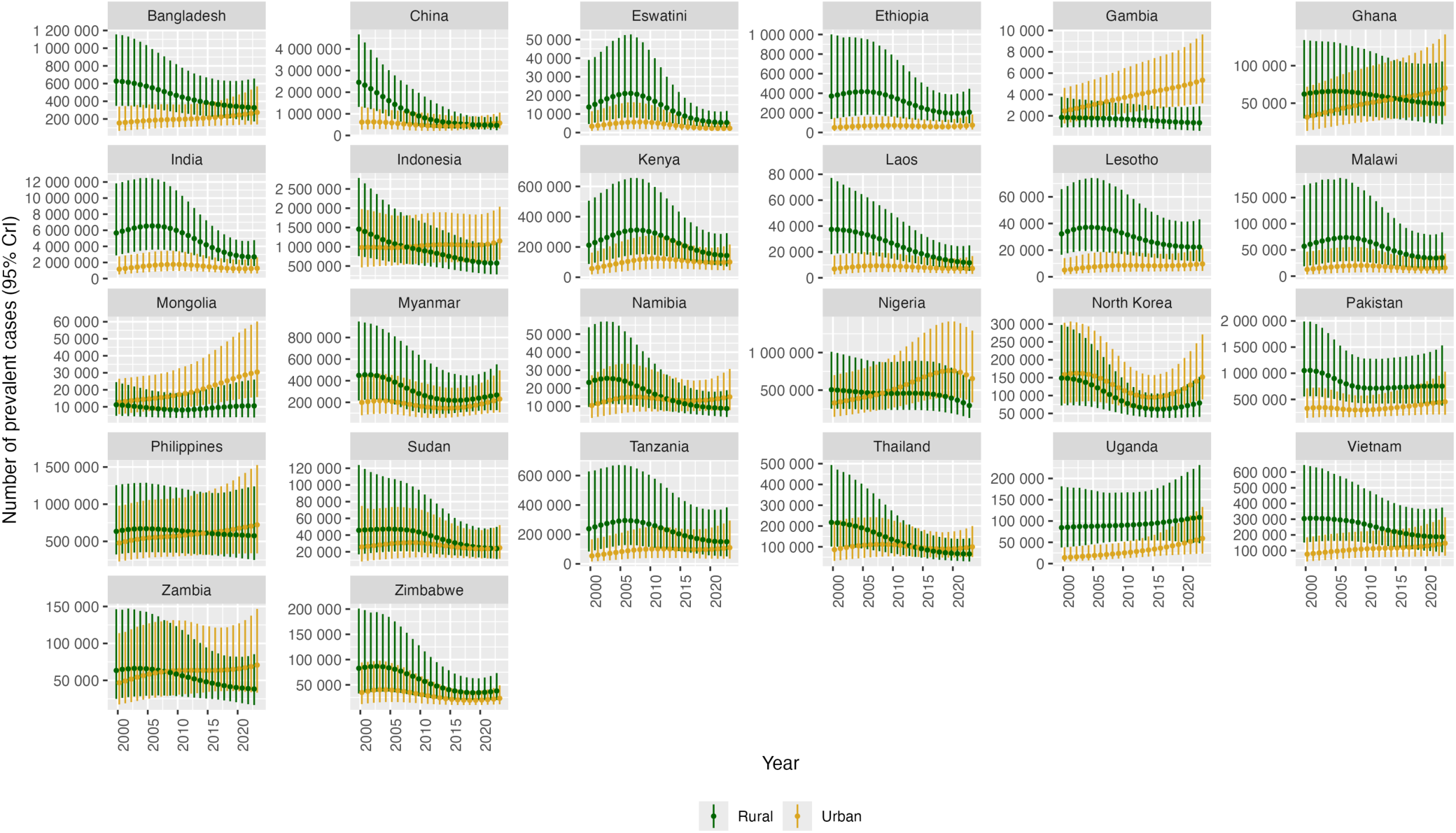
Estimated numbers of people with prevalent TB in urban and rural areas in 26 study countries between 2000 and 2023.

